# Multi-BOUNTI: Multi-lobe Brain vOlUmetry and segmeNtation for feTal and neonatal MRI

**DOI:** 10.64898/2026.04.21.26351376

**Authors:** Alena Uus, Abi Fukami-Gartner, Vanessa Kyriakopoulou, Daniel Cromb, Taeona Morgan, Sophie Arulkumaran, Alexia Egloff Collado, Aysha Luis, Roos Bos, Antonis Makropoulos, Andreas Schuh, Emma Robinson, Helena Sousa, Maria Deprez, Lucilio Cordero-Grande, Charline Bradshaw, Kathleen Colford, Jana Hutter, Anthony Price, Jonathan O’Muircheartaigh, Alexander Hammers, Daniel Rueckert, Serena Counsell, Grainne McAlonan, Tomoki Arichi, A. David Edwards, Joseph V. Hajnal, Mary A. Rutherford, Lisa Story

**Affiliations:** Research Department of Early Life Imaging, School of Biomedical Engineering and Imaging Sciences, King’s College London, UK; Department of Neonatology, UMC Utrecht, Utrecht, The Netherlands; Department of Computing, Imperial College London, London, UK; Research Department of Biomedical Computing, School of Biomedical Engineering and Imaging Sciences, King’s College London, London, UK; Biomedical Image Technologies, Universidad Politécnica de Madrid and CIBER-BBN, Madrid, Spain; Department of Forensic and Neurodevelopmental Science, School of Academic Psychiatry, Kings College London, London, UK; PET Imaging Centre, School of Biomedical Engineering and Imaging Sciences, King’s College London, London, UK; Chair for AI in Healthcare and Medicine, Technical University of Munich (TUM) and TUM University Hospital, Munich, Germany; Munich Center for Machine Learning (MCML), Munich, Germany; Research Department of Imaging Physics and Engineering, School of Biomedical Engineering and Imaging Sciences, King’s College London, London, UK; Institute for Information Processing, Leibniz University Hannover, Hannover, Germany; Department of Women and Children’s Health, School of Life Course and Population Sciences, King’s College London, London, UK; Fetal Medicine Unit, Guy’s and St Thomas’ NHS Foundation Trust, London, UK

**Author notes:** Corresponding author: Alena Uus; Joint senior authors: M.R. and L.S. *Email address:* (Alena Uus).

**Keywords:** Automated segmentation, Brain parcellation, Fetal brain MRI, Neonatal brain MRI, Normative growth modelling

## Abstract

Regional volumetric assessment of perinatal brain development is currently limited by the lack of consistent high quality multi-regional segmentation methods applicable to both fetal and neonatal MRI. We present Multi-BOUNTI, a deep learning pipeline for automated multi-lobe segmentation of fetal and neonatal T2w brain MRI. The method is based on a dedicated 43-label parcellation protocol and a 3D Attention UNet trained on brain MRI datasets of subjects spanning 21–44 weeks gestational/postmenstrual age. The pipeline integrates preprocessing, segmentation and volumetric analysis, and was evaluated on independent datasets, demonstrating fast (< 10 min/case) and accurate performance with high agreement to manually refined labels.

We demonstrate the application of the framework with 267 fetal and 593 neonatal MRI datasets from the developing Human Connectome Project without reported clinically significant brain anomalies to derive normative volumetric growth models across 21–44 weeks GA/PMA. These models were used to characterise developmental trajectories, assess differences between fetal and preterm neonatal cohorts, and analyse longitudinal changes. The resulting normative models were integrated into an automated reporting framework enabling subject-specific volumetric assessment via centiles and z-scores.

Multi-BOUNTI provides a unified and scalable approach for perinatal brain segmentation and volumetry, supporting large-scale studies and facilitating future clinical translation. The full pipeline is publicly available at https://github.com/SVRTK/perinatal-brain-mri-analysis.

## 1. Introduction

Automated segmentation of brain structures is a key component of modern fetal and neonatal magnetic resonance imaging (MRI) studies of brain development, supporting both exploratory research and clinical applications. Typically performed on structural T2-weighted (T2w) MRI, it provides regional volumetric measurements for normative modelling (Dimitrova et al., 2021; Cromb et al., 2024) and serves as an essential input for cortical surface analysis (Ma et al., 2025), quantitative MRI relaxometry (Payette et al., 2025b), as well as a key component of functional and diffusion MRI processing pipelines for connectomic analysis. Automated volumetric assessment of abnormal anatomy (Deprest et al., 2023) also has strong potential for integration into clinical radiological workflows, which still predominantly rely on manual 2D biometry of major brain structures. Consequently, both segmentation quality and the anatomical fidelity of parcellation protocols directly affect downstream analysis.

Introduced over a decade ago, classical registrationbased pipelines for 3D fetal and neonatal brain segmentation remain the predominant reference standard. The Draw-EM pipeline (Makropoulos et al., 2018), developed within the developing Human Connectome Project (dHCP) (Edwards et al., 2022), uses ALBERT atlas label propagation and expectation maximisation clustering, enabling the first standardised multi-regional (87 labels) segmentation of large fetal and neonatal cohorts and has been widely adopted. The more recently introduced M-CRIB-S pipeline for multi-lobe neonatal brain segmentation also relies on registration which is then followed by surface-based refinement of the cortical ribbon (Adamson et al., 2024). Similarly, the CRL fetal brain atlases with more than 100 regions (Gholipour et al., 2017; Bagheri et al., 2026) are widely used for label propagation and downstream analysis in many studies. However, these classical approaches are typically associated with prolonged runtime, overestimation of the cortical ribbon and reduced registration accuracy in low-quality images and atypical anatomy, such as preterm neonates or severe anomalies.

More recently, Draw-EM and CRL atlas label propagation segmentations have been used to train 3D deep learning (DL) models, such as 3D UNet (Cicek et al., 2016) or nnUNet (Isensee et al., 2021), for T2w fetal (Huang et al., 2026; Zeng et al., 2026; Bagheri et al., 2026) and neonatal (Grigorescu et al., 2021; Richter and Fetit, 2022; Henschel et al., 2024; Mihailov et al., 2025) MRI. Although these approaches offer substantially improved computational efficiency, errors and biases present in the training labels inherently propagate to the DL outputs. The FeTA fetal brain segmentation challenge (Payette et al., 2025a) has led to multiple methods, including approaches targeting abnormal brain segmentation with varying image quality and acquisition protocols (Deprest et al., 2023; Fidon et al., 2024; Zalevskyi et al., 2026); however, the associated parcellation protocol is limited to eight global regions of interest (ROIs), and the manual ground truth labels contain inaccuracies. The existing deep learning segmentation solutions for wider paediatric age range such as BIBSNet (Hendrickson et al., 2026) also tend to provide only global tissue parcellations. We have recently described the BOUNTI fetal brain DL segmentation pipeline (Uus et al., 2023) partially addresses these limitations by using high-quality manually refined labels for training, but remains restricted to 19 parcellation ROIs, lacks lobewise subdivision of grey and white matter, and is not applicable to neonatal MRI.

To the best of our knowledge, no existing DL-based methods provide high-quality multi-lobe detailed segmentation applicable to both fetal and neonatal brain MRI. This limitation is particularly relevant given the need for harmonised analysis across the perinatal period to enable continuous modelling of developmental trajectories (Urru et al., 2025). Furthermore, including more regions may lead to increased understanding of brain development.

### 1.1. Contributions

Here we address the need for a harmonised multi-lobe segmentation tool by introducing a prototype deep learning framework for combined fetal and neonatal multi-lobe brain segmentation of structural 3D T2w MRI.

The approach is based on a dedicated 43-label parcellation protocol specifically designed to ensure clinical and research relevance across the perinatal period, from the late second trimester to postnatal stages. The pipeline integrates preprocessing with a 3D Attention UNet trained on 160 fetal and neonatal scans and evaluated on 40 datasets, with qualitative comparison to existing multilobe segmentation methods.

Next, we demonstrate a key application of our approach by deriving normative perinatal volumetric growth models across 21–44 weeks gestational (GA) and postmenstrual age (PMA) from 267 fetal and 593 neonatal (term and preterm) dHCP datasets without reported clinically significant brain findings. These models are integrated into an automated reporting framework for subjectspecific volumetric assessment, providing percentiles and z-scores in .html format for potential clinical and research applications.

## 2. Methods

### 2.1. Cohorts and datasets

This study uses T2-weighted fetal and neonatal brain MRI datasets from the dHCP project (REC:14/LO/1169), acquired on a 3T Philips Achieva system (Philips Healthcare, Netherlands) using dedicated protocols and reconstructed to 0.5 mm isotropic resolution in standard radiological space (Price et al., 2019; Cordero-grande et al., 2019; Edwards et al., 2022). All datasets were reviewed and radiologically assessed by an experienced perinatal neuroradiologist (MR).

The full cohort used for training, testing, analysis and normative modelling comprised 274 fetal (146 males; scan GA: 29.13 ± 3.94, range 20.86–38.29 weeks) and 636 neonatal (343 males; scan PMA: 40.00 ± 3.70, range 26.71–45.14 weeks; birth GA: 38.15 ± 4.29, range 23–43 weeks) datasets. The general inclusion criteria were sufficient image quality with clear visibility of key brain structures and absence of severe signal or motion artefacts (Uus et al., 2022; Cuadra et al., 2025). The training and testing cohort (200) included both control and abnormal datasets. For the normative growth modelling and analysis (860), we used only the cases without reported clinically significant brain anomalies. The training/testing and normative modelling cohorts were not mutually exclusive, with eligible datasets contributing to both analyses.

In addition, as part of an extended generalisability experiment, we used 405 fetal brain MRI reconstructions from the iFIND project (REC: 14/LO/1806). The T2w SSTSE datasets were acquired on a 1.5T Philips Ingenia system (Philips Healthcare, Netherlands), with TE = 80 ms, an in-plane resolution of 1.25×1.25 mm, 2.5 mm slice thickness and -1.25 mm gap, and reconstructed using the auto-SVRTK method (Kuklisova-Murgasova et al., 2012; Uus et al., 2025) in standard radiological space before resampling to 0.5 mm isotropic resolution.

### 2.2. Multi-lobe brain parcellation protocol

The multi-regional parcellation protocol was codesigned by neuroradiologists and imaging scientists (AFG, VK, AU, DC, RB, SA, AL, MR) through iterative discussions, consensus meetings and literature review. It builds on the ALBERT atlas (Gousias et al., 2012) from Draw-EM (87 labels) (Makropoulos et al., 2018) and BOUNTI (19 labels) (Uus et al., 2023) protocols, while maintaining compatibility with established neuroanatomical atlases by Bayer and Altman (Bayer and Altman, 2003, 2005). Additional reference was made to parcellation schemes from existing tools, including M-CRIBS (Adamson et al., 2024) and CRL atlas (Bagheri et al., 2026).

Region selection was guided by clinical and research relevance, as well as the visibility of ROI interfaces and anatomical landmarks on T2w contrast across the 21–44 weeks age range. Due to rapid developmental changes and variable image quality, certain anatomical boundaries (e.g., major sulci before 24 weeks GA or the internal capsule separating deep grey matter structures) may not be well defined due to low visibility. Where required for downstream analysis, these interfaces were approximated based on expected anatomical location. The protocol was defined using atlas label propagation followed by manual sub-parcellation and refinement in 4 fetal and 4 neonatal datasets spanning 21–44 weeks GA using ITK-SNAP (Yushkevich et al., 2006).

The final protocol comprises 43 labels. Cortical grey matter (GM) and white matter (WM) are subdivided into six major lobar ROIs per hemisphere (frontal, parietal, temporal, occipital, insular and cingulate). Deep GM is partitioned into three regions per hemisphere (caudate nucleus, basal ganglia and thalami). The intracranial cerebrospinal fluid (CSF) compartment includes perhemisphere extracerebral CSF and lateral ventricles, as well as the cavum (combined cavum septum pellucidum and vergae) and the third and fourth ventricles. The posterior fossa is divided into brainstem, cerebellar hemispheres and vermis, and the corpus callosum is segmented as a single label. As in Draw-EM, an additional intracerebral background label captures subcortical/diencephalic structures not explicitly delineated (e.g., internal capsule, hypothalamus, hippocampus), which are planned for further subdivision in future protocol extensions.

### 2.3. Multi-lobe brain segmentation pipeline

The proposed unified pipeline for automated deep learning-based multi-regional segmentation of 3D T2w fetal and neonatal brain MRI is summarised in Fig. 1.

**Figure 1:**
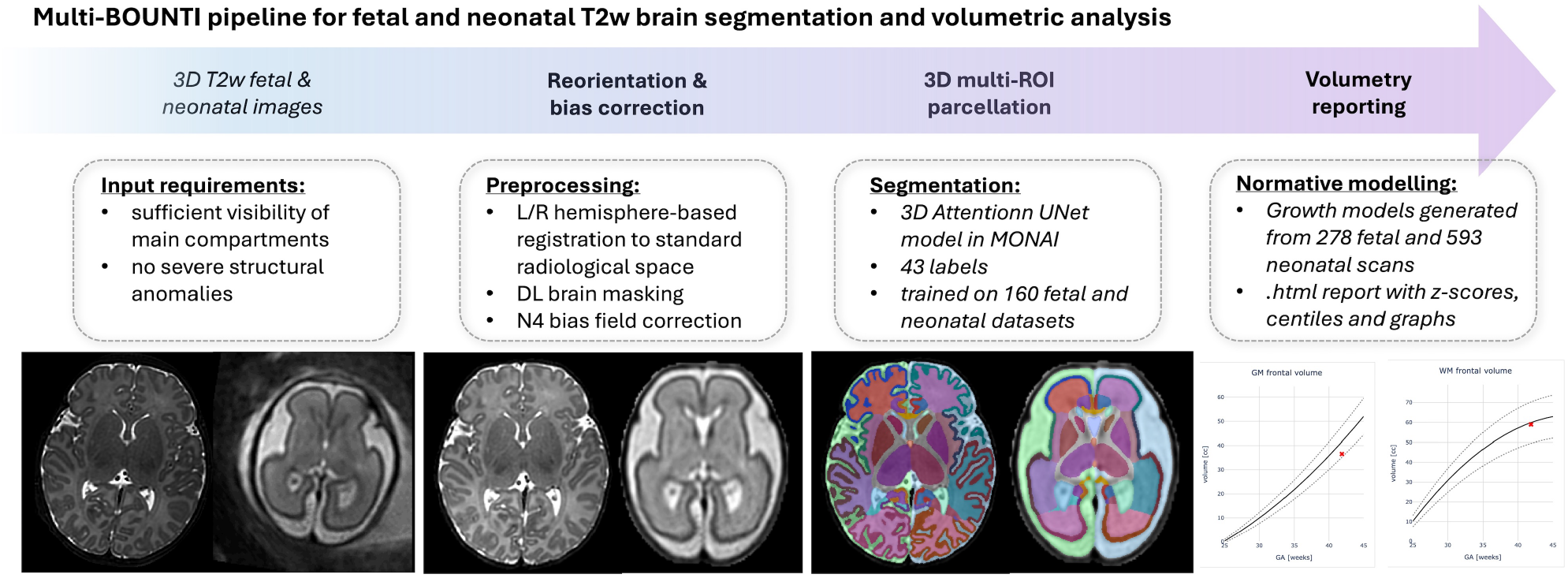
Proposed pipeline for multi-regional brain segmentation and volumetry in 3D T2w perinatal MRI.

As a preprocessing step, external background (i.e., scalp and surrounding intrauterine structures) is removed via automated brain extraction using a pretrained in-house fetal and neonatal pipeline (Uus et al., 2023) based on a 3D Attention UNet (Oktay et al., 2018). Since lobe-wise parcellation is driven by cortical morphology and brain proportions, segmentation performance is sensitive to image orientation, which varies across subjects. Therefore, alignment to standard radiological space is performed to ensure consistent segmentation. This is achieved via affine registration of left and right (L/R) hemisphere labels to an atlas, correcting for pose, scale and shear. Hemisphere labels are generated using a 2-label 3D Unet (Cicek et al., 2016) trained on 80 fetal and 80 neonatal datasets with labels derived from BOUNTI (Uus et al., 2023). Registration is implemented using MIRTK^1^. The input image is then transformed to atlas space, followed by masking, N4 bias field correction (Tustison and Gee, 2009), and resampling to a 256 × 256 × 256 grid.

Similarly to our previous work (Uus et al., 2023), the main segmentation network employs a classical 3D Attention UNet with five encoder–decoder blocks (channels: 32, 64, 128, 256, 512), kernel size 3, ReLU activation, dropout 0.5, and batch size of one. Training uses the AdamW optimiser with a linearly decaying learning rate (initialised at 1 × 10^−4^), default β parameters, and weight decay 1 × 10^−5^. Optimisation is performed with combined Dice and cross-entropy loss over 200,000 iterations. Data augmentation includes intensity normalisation, random cropping (128 × 128 × 128), bias field simulation, sagittal flipping, and ±5^°^ rotations. The training set comprises 90 fetal and 70 neonatal datasets from 21–44 weeks GA/PMA range. This cohort includes both typical and abnormal cases to facilitate robust performance for anatomical variations. Ground truth labels were generated using an iterative active learning approach, combining network pretraining, inference on new cases, manual refinement, and retraining. All refinements were performed in ITKSNAP by a researcher (AU) with more than 7 years of experience in fetal and neonatal MRI.

Predicted segmentations are transformed back to native space for downstream analysis. Regional volumetry (cc) is computed for all labels, alongside lobe-specific cortical surface area and global gyrification index (GI), defined as the ratio of cortical surface area to a morphologically closed outer surface (Zilles et al., 1988).

The pipeline is implemented as a unified bash script with a simple interface. Deep learning components are developed within MONAI framework (Cardoso et al., 2022). Source code and trained models are publicly available (https://github.com/SVRTK/perinatal-brain-mri-analysis), with deployment provided via a standalone Docker supporting CPU and GPU execution.

Given the impracticality of fully manual delineation of all 43 regions, quantitative evaluation was performed against manually refined automated segmentations. As these reference labels were derived from model predictions, this assessment primarily measures agreement with manual refinement rather than fully independent segmentation accuracy. The test set includes 40 independent fetal (20) and neonatal (20) datasets selected to cover the whole 21–45 weeks GA/PMA range. Segmentations were visually inspected and refined in ITK-SNAP (AU), and all 43 labels were evaluated using Dice scores and relative volumetric differences. Qualitative scoring per ROI was also performed (4: good; 3: acceptable, minor imperfections; 2: suboptimal, significant errors; 1: poor, failed segmentation).

A direct quantitative comparison with existing methods across all regions is not feasible because their parcellation protocols differ and no common ground truth exists. We therefore performed a qualitative comparison on six datasets (three fetal, three neonatal) against eight publicly available multi-regional segmentation methods with open-source implementations.

The four neonatal methods comprised two classical registration-guided approaches, Draw-EM (Makropoulos et al., 2018) (87 labels) and M-CRIB-S (Adamson et al., 2024) (101 labels), and two deep learning methods. VINNA (Henschel et al., 2024) is a network trained on dHCP neonatal data with Draw-EM-derived parcellations (87 labels). BIBSNet (Hendrickson et al., 2026) is an nnU-Net trained on infant data (0–8 months) (35 labels).

The four fetal methods comprised two classical registration-guided approaches, Draw-EM (Makropoulos et al., 2018) (91 labels) and label propagation from the CRL fetal brain atlases (Gholipour et al., 2017) (32 labels), and two deep learning methods. Trustworthy-AI (TW-AI) (Fidon et al., 2024) is an ensemble of five 3D nnU-Nets (8 labels). FetalSynthSeg (Zalevskyi et al., 2026) is a 3D U-Net trained on synthetic and real FeTA data (7 labels).

In addition, we quantitatively assessed cortical grey matter segmentation, the one label defined consistently across all protocols. For each subject, cortical GM was manually delineated on a single representative mid-axial slice in ITK-SNAP by one rater (AU). Each method’s output, including Multi-BOUNTI, was restricted to the same slice and binarised to cortical GM. It was then compared with the manual reference using the Dice similarity coefficient, the 95th percentile Hausdorff distance, the average symmetric surface distance, and the relative volume differences. All metrics were computed in 2D within the annotated slice.

### 2.4. Normative growth modelling

To assess the utility of the proposed pipeline for largescale analysis and normative modelling, it was applied to a selected cohort of 267 fetal, 494 term, and 99 preterm neonatal 3T T2w datasets from the dHCP project. Inclusion criteria for fetal and term neonatal controls were: singleton pregnancy, sufficient image quality, acceptable segmentation quality, absence of clinically significant brain anomalies, and >5th birth weight centile. For preterm neonates, criteria included sufficient image quality, acceptable segmentation quality, absence of clinically significant anomalies, and >5th birth weight centile. All segmentations underwent visual quality control (AU, AFG). Extracted regional volumetric and cortical metrics (left/right hemisphere structures were combined) were used to characterise developmental trajectories across the perinatal period. Statistical analysis was performed in Python using statsmodels. Analysis of covariance (ANCOVA) assessed associations with age at scan, sex, parental ethnicity, birth weight centile, and mode of delivery (neonates), as well as group differences between fetal and preterm neonatal cohorts. Developmental effects were modelled using linear and quadratic terms of GA and PMA at scan. Multiple comparisons across ROIs were controlled using Bonferroni correction.

Normative growth models were then derived to enable subject-specific estimation of centiles and z-scores. Model fitting was performed separately for fetal and neonatal cohorts, with the latter including both term and preterm neonates to ensure continuity across the age range. The 5th, 50th, and 95th centiles were estimated using polynomial regression (NumPy/SciPy), following established perinatal modelling approaches (Royston and Wright, 1998).

Finally, longitudinal analysis was performed in a subset of 10 subjects with three or four fetal and neonatal scan timepoints. Growth rates were estimated using finite differences (Δ*volume*/Δ*age*) between consecutive scans, where age denotes GA for fetal and PMA for neonatal scans, and summarised across subjects.

### 2.5. Automated reporting

Integration of the proposed segmentation pipeline into clinical workflows requires an intuitive and interpretable output format to support case-specific analysis.

Following segmentation, the 3D T2w image and corresponding segmentation are passed to a dedicated Python script (fetal or neonatal), which automatically generates an .html report. The report includes:

- general case information (patient identifier, scan date, GA/PMA);
- 21 representative T2w images with segmentation overlays (axial, coronal and sagittal views);
- tabulated measurements with observed-to-expected ratios, centiles and z-scores;
- growth charts comparing extracted measurements to normative centile curves;
- a disclaimer indicating research-only use pending regulatory approval for clinical deployment.

### 2.6. Initial evaluation of generalisation and anomaly handling

To explore the generalisability of Multi-BOUNTI beyond its training data, we performed two additional qualitative evaluations. First, the pipeline was applied to an independent iFIND data release cohort comprising 405 reconstructed fetal brain MRI datasets acquired at 1.5T, including normal and abnormal cases, predominantly with cardiovascular anomalies, and varying image quality. All segmentations were visually inspected (by AU) and assigned a quality grade based on the presence and severity of segmentation errors: (1) severe errors, (2) significant errors, (3) minor errors, or (4) good quality. This assessment provided an initial evaluation of generalisation to different acquisition protocols and anatomical presentations, given that the pipeline was trained predominantly on normal anatomy from the dHCP 3T acquisition protocol.

Second, we presented representative segmentation examples for fetal and neonatal datasets with reported clinically significant anomaly findings to illustrate the method’s performance in the presence of abnormal anatomy. These examples included nine fetal and neonatal T2w reconstructions from the dHCP and iFIND cohorts (excluded from training) and three abnormal fetal brain MRI reconstructions from the FeTA challenge cohort (Payette et al., 2025a).

For the global fetal brain extraction and masking of non-dHCP 1.5T fetal datasets, we utilized the network weights from the original BOUNTI pipeline (Uus et al., 2023).

## 3. Results

### 3.1. Multi-regional brain parcellation protocol

The final Multi-BOUNTI parcellation protocol with 43 labels is summarised in Fig. 2, alongside examples of 3D DL segmentation outputs across fetal and neonatal scan time points (22–44 weeks GA/PMA). As expected, pronounced age-related changes in cortical folding are observed, together with gradual shifts in the relative proportions of the cerebellum and lateral ventricles. Notably, cortical GM and WM exhibit similar visual appearance at overlapping fetal and neonatal time points (e.g., 30 and 38 weeks).

**Figure 2:**
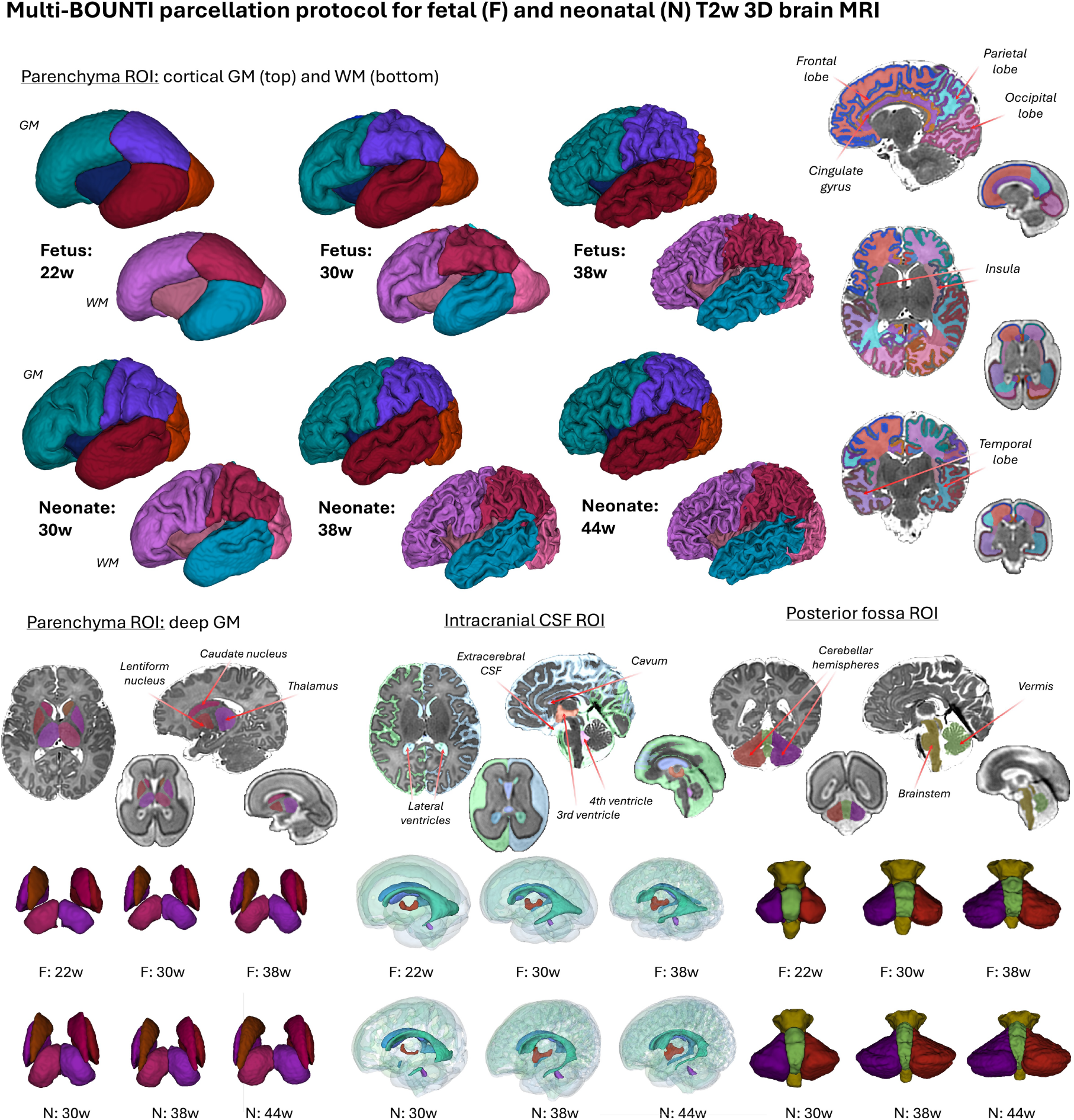
Proposed multi-BOUNTI parcellation protocol for fetal and neonatal 3D T2w brain MRI with DL segmentation examples at different time points.

During protocol development, particular emphasis was placed on accurate delineation of cortical folds and exclusion of small CSF pockets and vascular regions, which are commonly misclassified due to similar intensity and partial volume effects in the existing methods. The resulting parcellation provides a consistent global anatomical framework that can be further extended, including finer sub-parcellation of lobes and intracerebral background structures.

### 3.2. Multi-lobe brain segmentation pipeline

Evaluation on 20 fetal and 20 neonatal independent datasets (21–45 weeks age at scan), summarised in Fig. 3, demonstrates the feasibility and efficiency of the proposed pipeline. Full processing requires less than 10 minutes per case on a standard engineering workstation (hardware requirements: > 16GB GPU). Per-label qualitative scores were predominantly in the 3–4 range (acceptable–good), with 142 (8.26%) minor imperfections (grade 3) and 8 (0.47%) significant or severe errors (grades 1–2) across 1,720 case-specific label assessments (40 datasets x 43 labels). These occurred mainly in cortical GM (frontal, parietal, temporal, occipital), cerebellar hemispheres, lateral ventricles and extracerebral CSF, primarily due to reduced contrast, limited regional visibility or vascular signal. Despite these local inaccuracies, the predicted segmentations showed high agreement with manually refined labels, with small relative volumetric differences and high Dice scores across most regions. Manual corrections required less than 2 minutes per case.

**Figure 3:**
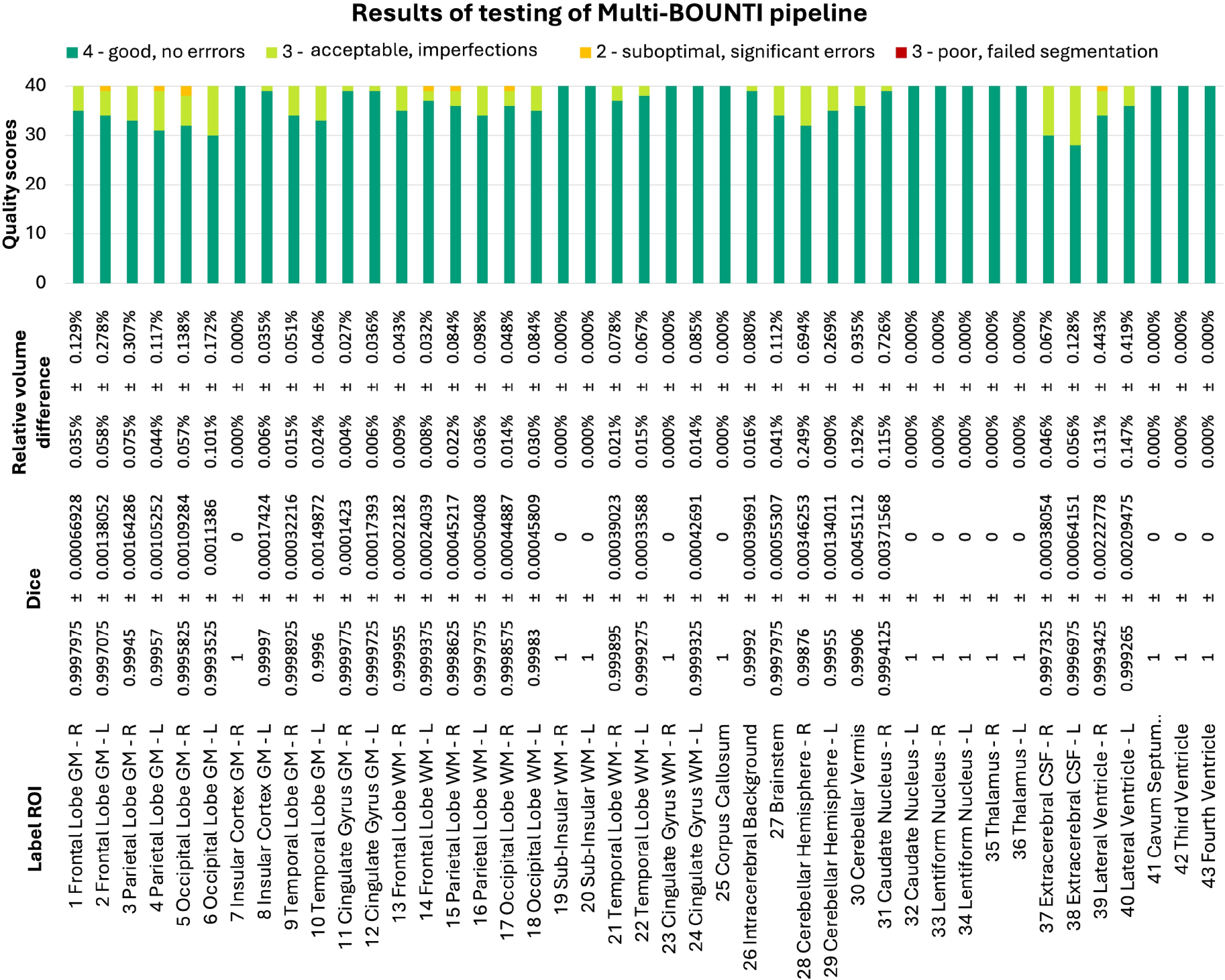
Summary results of testing on 20 fetal and 20 neonatal datasets: quality scores, relative volume difference and Dice per label.

Fig. 4 shows qualitative comparisons with publicly available multi-regional segmentation methods for neonatal (a) and fetal (b) T2w brain MRI. Across both populations, Multi-BOUNTI demonstrated more consistent anatomical delineation across ROIs, particularly around the ventricular system, deep grey matter and cortical boundaries. Differences were especially apparent in cortical grey matter, where several existing methods showed overestimation of the cortical ribbon, inclusion of adjacent white matter or CSF, and less accurate delineation of cortical folding. In contrast, Multi-BOUNTI provided a thinner and more anatomically consistent cortical ribbon, with improved correspondence to the underlying tissue boundaries. These observations were consistent across the representative developmental stages, suggesting improved robustness to the substantial changes in brain morphology and cortical maturation across the fetal–neonatal period.

**Figure 4:**
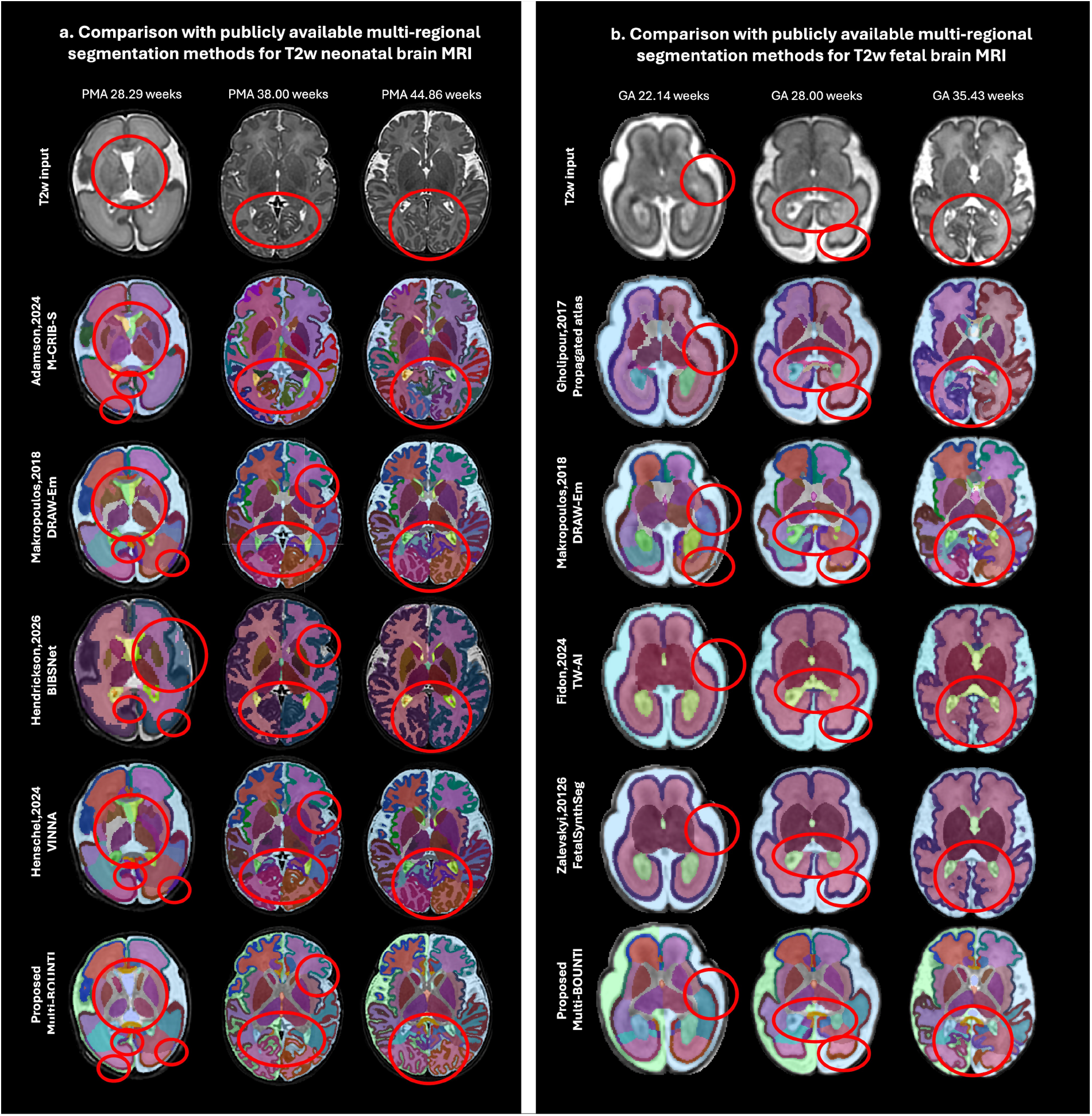
Qualitative comparison of Multi-BOUNTI segmentations with 8 other publicly-available neonatal (a) and fetal (b) multi-ROI parcellation methods. The regions with improved performance in Multi-BOUNTI outputs are highlighted in red. (Note: the label colours were adapted to be similar to the Multi-BOUNTI colourmap).

**Figure 5:**
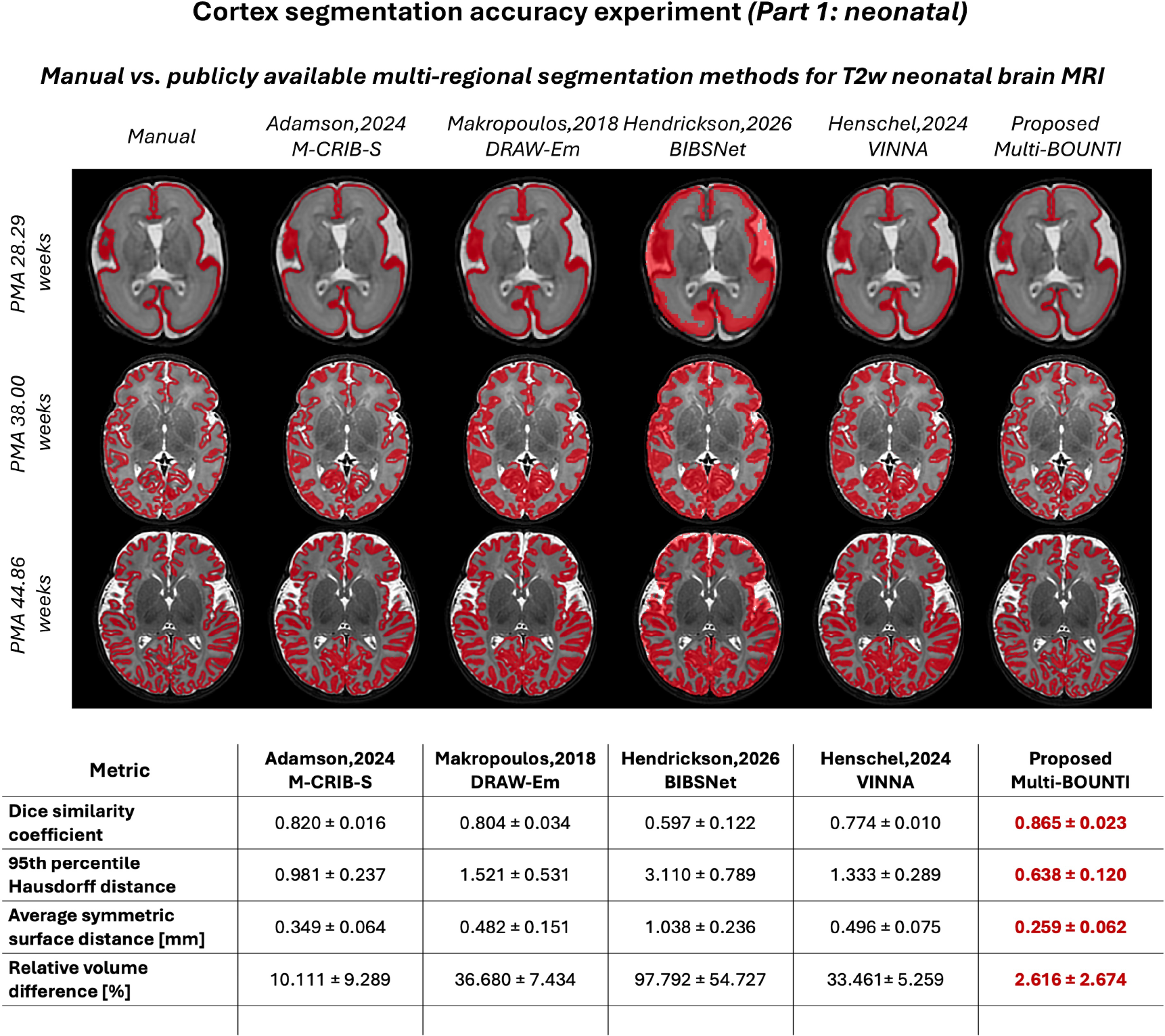
Quantitative comparison of manual vs. automated cortex segmentation in three neonatal datasets: Multi-BOUNTI and four other publicly available neonatal multi-ROI parcellation methods.

**Figure 6:**
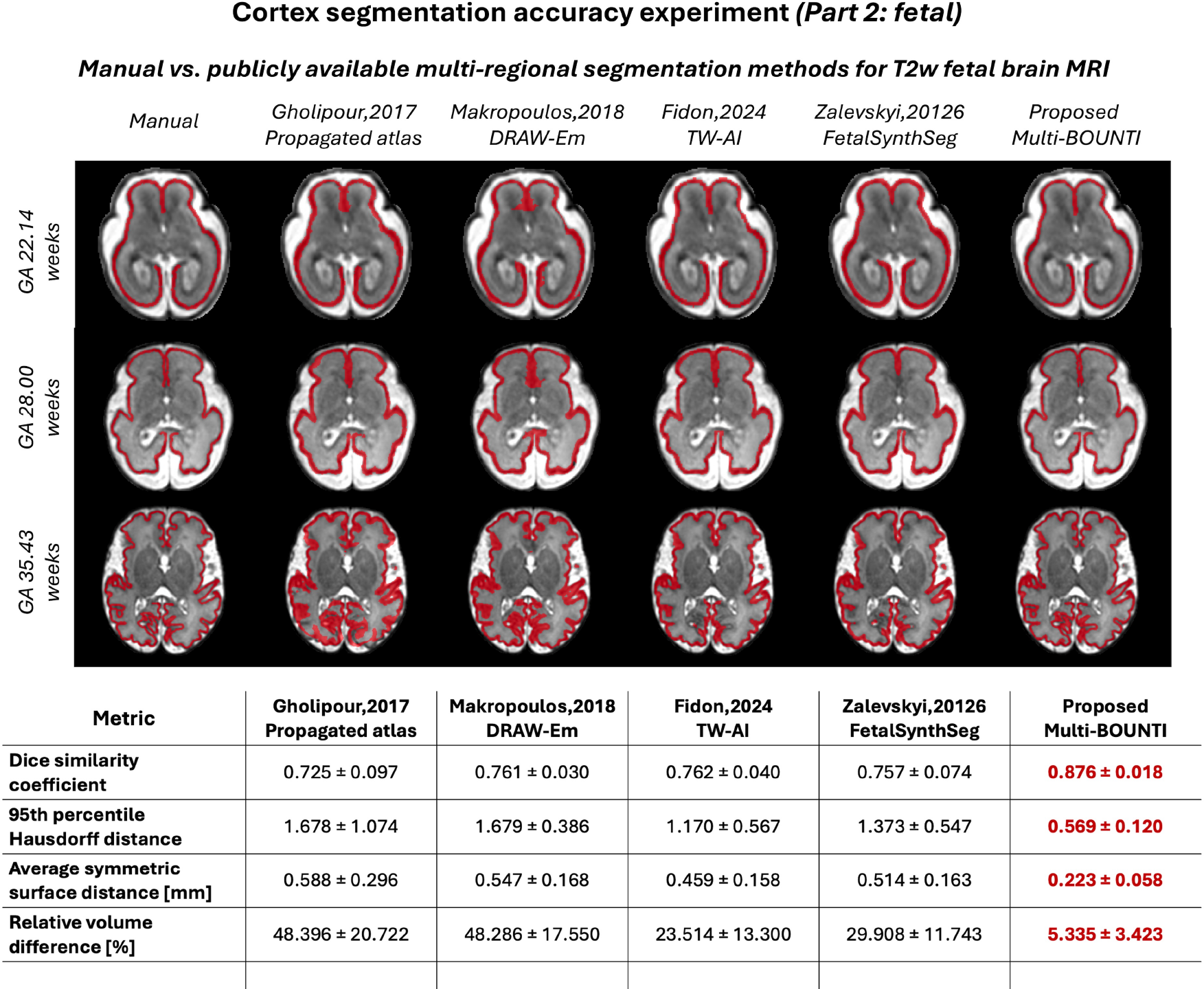
Quantitative comparison of manual vs. automated cortex segmentation in three fetal datasets: Multi-BOUNTI and four other publicly available multi-ROI parcellation methods.

This was further supported by the quantitative cortical GM segmentation experiment (Fig.5-6). Multi-BOUNTI achieved the best performance across the evaluated cortical GM segmentation metrics in the three fetal and three neonatal test cases. For neonatal MRI, it achieved a Dice coefficient of 0.865 ± 0.023, HD95 of 0.638 ± 0.120, ASSD of 0.259 ± 0.062 mm and relative volume difference of 2.616 ± 2.674%, outperforming all four comparison methods. Similarly, for fetal MRI, Multi-BOUNTI achieved a Dice coefficient of 0.876 ± 0.018, HD95 of 0.569 ± 0.120, ASSD of 0.223 ± 0.058 mm and relative volume difference of 5.335 ± 3.423%, again providing the best results across all metrics. The improvements in both volumetric overlap and surface-based measures indicate more accurate cortical boundary delineation, while the substantially lower volume differences suggest reduced overand under-segmentation. Together, these findings provide quantitative evidence supporting the improved cortical delineation observed visually in the evaluated fetal and neonatal datasets.

### 3.3. Normative growth modelling

The Multi-BOUNTI pipeline was applied to 267 fetal and 494 term and 99 preterm neonatal dHCP datasets without reported clinically significant brain anomalies (Fig. 7). After visual inspection all segmentations were confirmed to be suitable for downstream analysis. Global volumetric growth charts and corresponding derived normative models are shown in Fig. 8.

**Figure 7:**
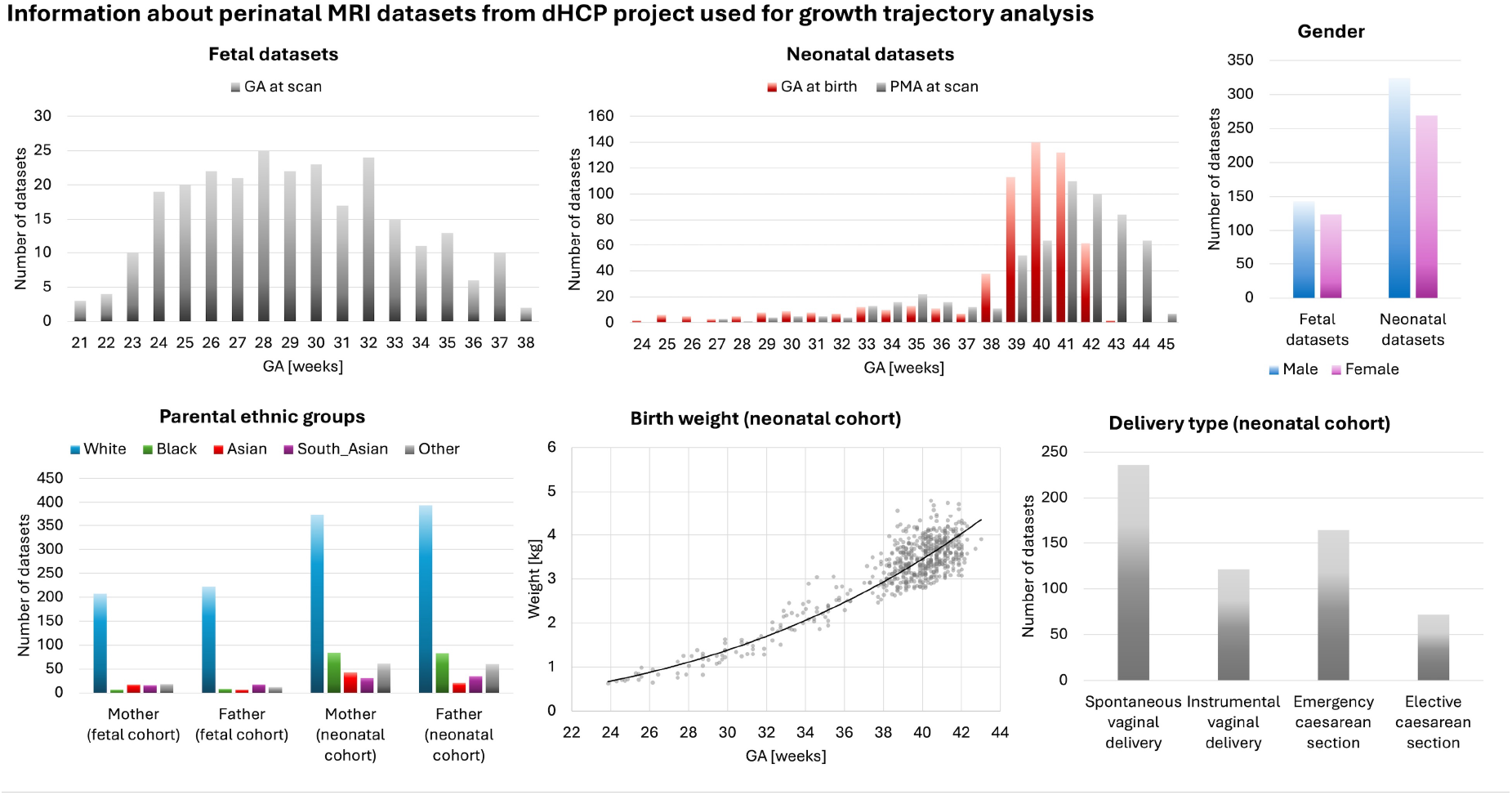
Summary information about dHCP datasets used for growth trajectory analysis and normative modelling (267 fetal and 593 neonatal).

**Figure 8:**
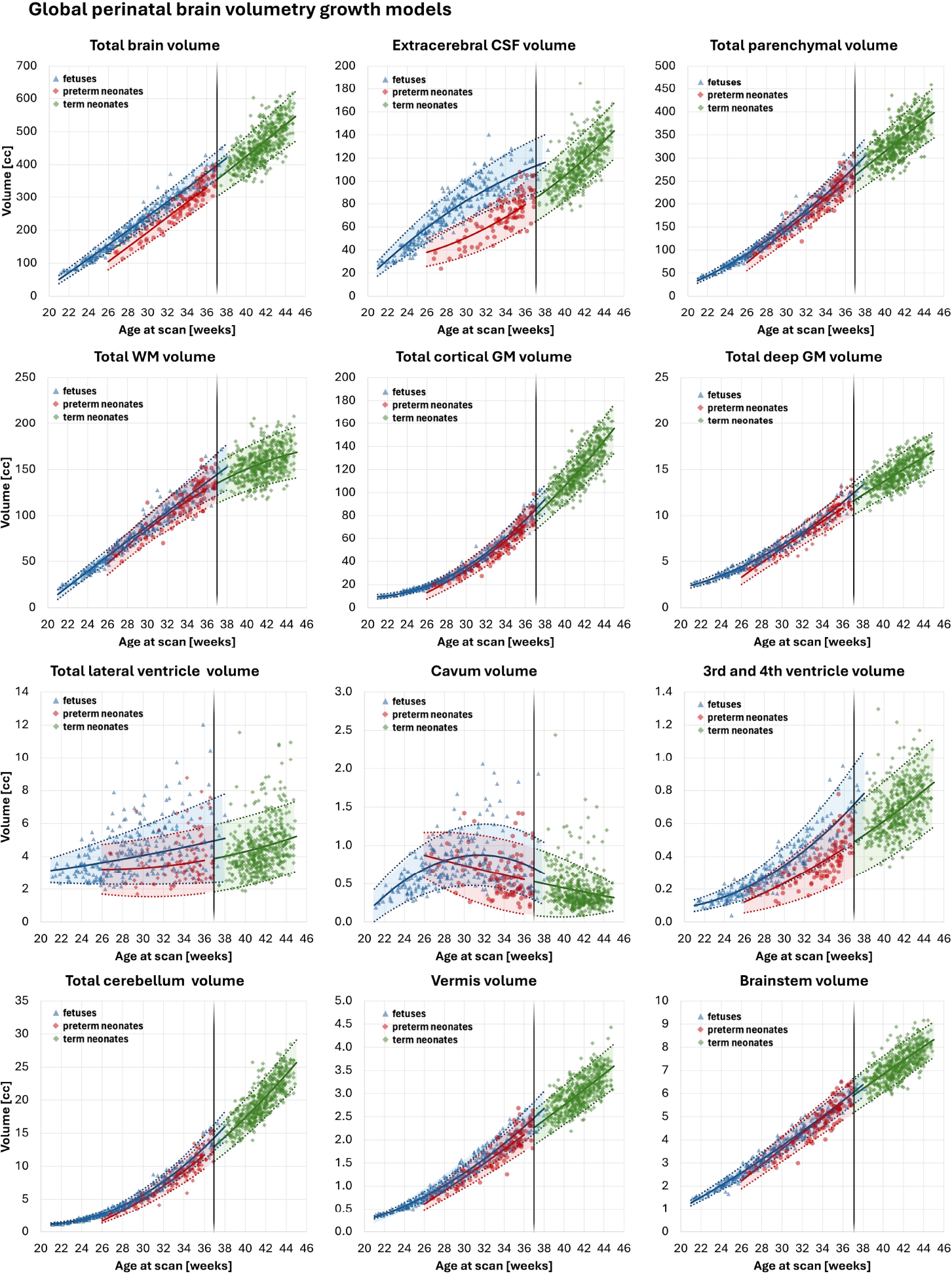
Global brain volumetry growth models of the fetal (267, blue) and term (494, green) and preterm (99, red) neonatal cohorts with derived 50th, 5th and 95th centiles. Note: these graphs include preterm cohort.

All structures followed expected developmental trajectories with clear inter-subject variability consistent with previous reports for the dHCP cohort (Dimitrova et al., 2021; Mihailov et al., 2025; Khan et al., 2026). Across both prenatal and postnatal cohorts, volumetric measures showed strong associations with age at scan, which was a highly significant predictor across all compartments (*p* < 0.0001). Parenchymal regions (total brain, parenchyma, cortical GM, WM, deep GM, cerebellum, vermis and brainstem) increased steadily with gestation and transitioned smoothly into the neonatal period beyond 37 weeks. Cortical GM exhibited nonlinear acceleration in the late third trimester, whereas WM and deep GM followed more gradual trajectories. In contrast, although all CSF compartments were significantly associated with age at scan they showed distinct patterns: extracerebral CSF and lateral ventricles were highly variable, cavum volume decreased with gestation, and third/fourth ventricles increased with GA.

In neonates, sex and birth weight centile were significantly associated with most regions (predominantly *p* < 0.0001), while delivery mode was not significant after correction. In fetuses, sex effects were limited to isolated regions (*p* < 0.01–0.05). No consistent associations with parental ethnicity were observed after Bonferroni correction, although interpretation is limited by imbalance in ethnic representation across cohorts.

Comparison between fetal and preterm neonatal cohorts within the overlapping period (26–36 weeks) revealed significant differences across most structures after correction for age at scan. Preterm neonates exhibited reduced parenchymal and CSF volumes compared to fetuses at equivalent age. Total brain volume showed the strongest effect (*p* < 0.0001), with similarly robust differences in parenchymal compartments and lobar volumes (all *p* < 0.0001). CSF compartments showed pronounced divergence, particularly in extracerebral CSF (*p* < 0.0001) with markedly smaller volumes in the preterm cohort, while cavum and ventricular effects were weaker. These differences may partly reflect early postnatal physiological adaptation (e.g., transient extracellular fluid contraction, diuresis, altered CSF dynamics and cranial moulding) (Modi et al., 2000; National Guideline Alliance (UK), 2017). Various antenatal and postnatal factors associated with preterm birth reported in other works (Ball et al., 2017; Story et al., 2021; Dimitrova et al., 2021) also potentially contribute to the lower volumes in the preterm cohort. The apparent discontinuity in lateral ventricular volumes between fetal and neonatal cohorts should be interpreted cautiously, as these represent different populations and acquisition contexts rather than longitudinal changes at birth. Furthermore, differences in cortical GM volumetry (*p* < 0.0001) may also partly reflect technical factors such as contrast, signal-to-noise ratio, and partial-volume effects, with intrinsically lower fetal image resolution and super-resolution reconstruction potentially contributing to GM overestimation.

Collectively, these results demonstrate differences in brain volumes between fetal and preterm neonatal cohorts at equivalent developmental ages, highlighting the impor- tance of accounting for acquisition context (in utero vs. ex utero) when modelling volumetric development across 21—4 weeks.

Lobe-specific trajectories (Fig. 9) showed consistent and regionally distinct maturation patterns. Cortical GM and WM volumes increased consistently across longitudinal timepoints, following subject-specific trajectories broadly aligned with the cross-sectional normative curves. Frontal and parietal lobes had the largest volumes, followed by temporal and occipital regions, while insular and cingulate regions remained smaller. Relative scaling between lobes remained stable, indicating coordinated growth. Cortical surface area closely followed GM trajectories and prior reports (Xu et al., 2022), with rapid expansion across all lobes, particularly in frontal and parietal regions. The gyrification index showed a nonlinear increase, with minimal change before 28–30 weeks and accelerated growth thereafter consistent with previous findings on the rapid increase in gyrification towards term (Lefèvre et al., 2016; Mihailov et al., 2025). This was most pronounced in occipital and parietal cortices suggesting a posterior-to-anterior gradient of cortical folding (Orasanu et al., 2016), while frontal and temporal regions showed later but substantial increases. In comparison, insular and cingulate regions exhibited smaller GI ranges. These results indicate preserved spatial organisation alongside region-specific differences in growth dynamics.

**Figure 9:**
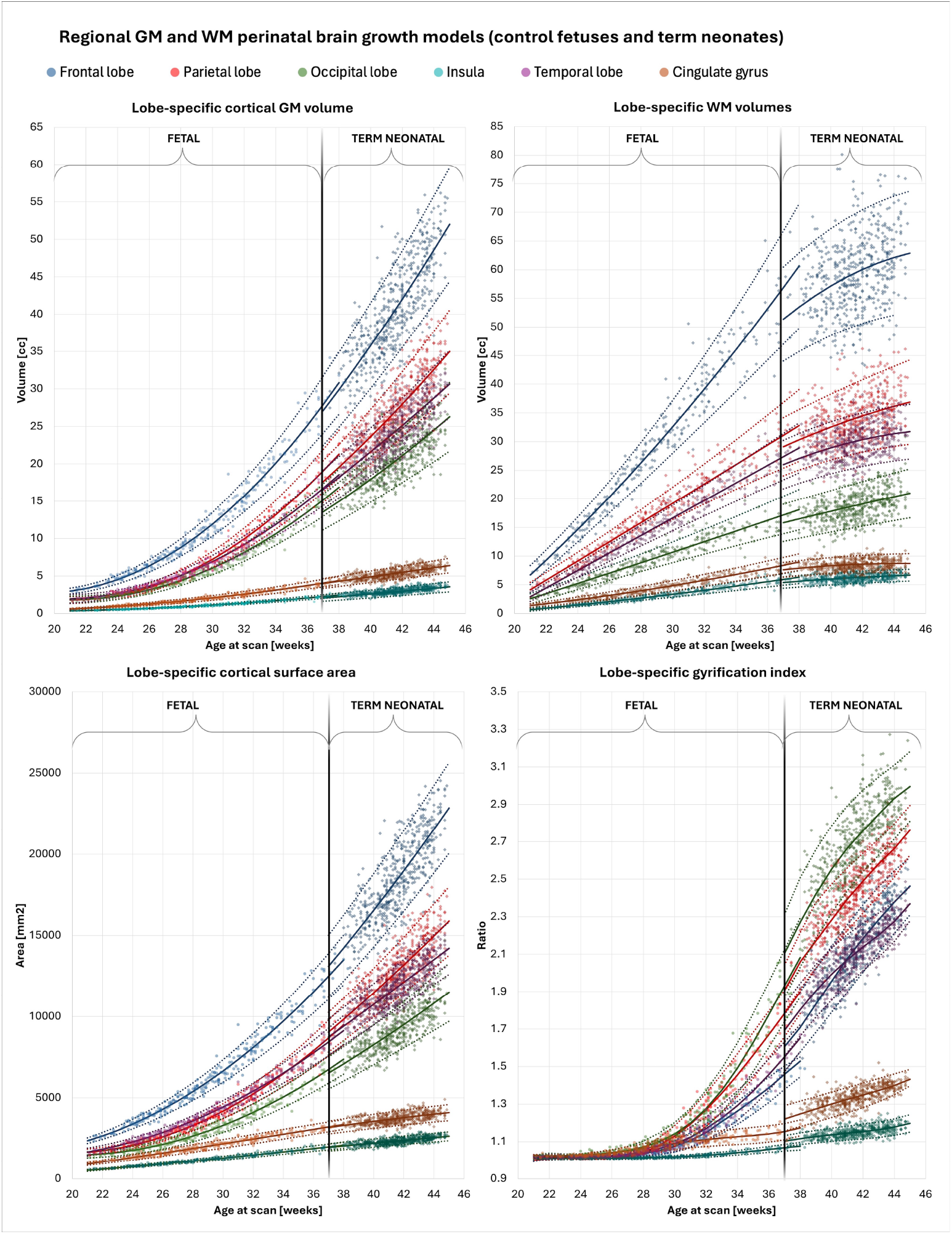
Lobe-specific cortical GM and WM growth models of the fetal (267) and term neonatal (494) cohorts (excluding preterm neonates) with derived 50th, 5th and 95th centiles. Note: these graphs do not include preterm cohort.

Longitudinal analysis of 10 subjects with multiple timepoints (Fig. 10) showed trajectories consistent with cross-sectional models. Cortical GM and WM volumes increased consistently across longitudinal timepoints, following subject-specific trajectories broadly aligned with the cross-sectional normative curves. Centile values were largely preserved within similar ranges, with modest within-subject variation. Growth rates were regionspecific, with highest in frontal GM (∼2.05 cc/week), followed by parietal (∼1.42), temporal (∼1.22), and occipital (∼1.02), while subcortical structures grew more slowly (e.g., thalamus ∼0.34), and cerebellar growth remained relatively high (∼1.02). In contrast, CSF compartments showed more variable dynamics. Extracerebral CSF volume generally increased with age, although growth rates were more variable, while lateral ventricles exhibited low growth rates (∼0.07 cc/week) and bidirectional changes. Cavum volume remained largely stable or decreased (0.01 cc/week), consistent with involution. Overall, parenchymal growth was consistent and generally accelerated with age with preserved scaling, whereas fluid compartments showed greater variability and less structured trajectories.

**Figure 10:**
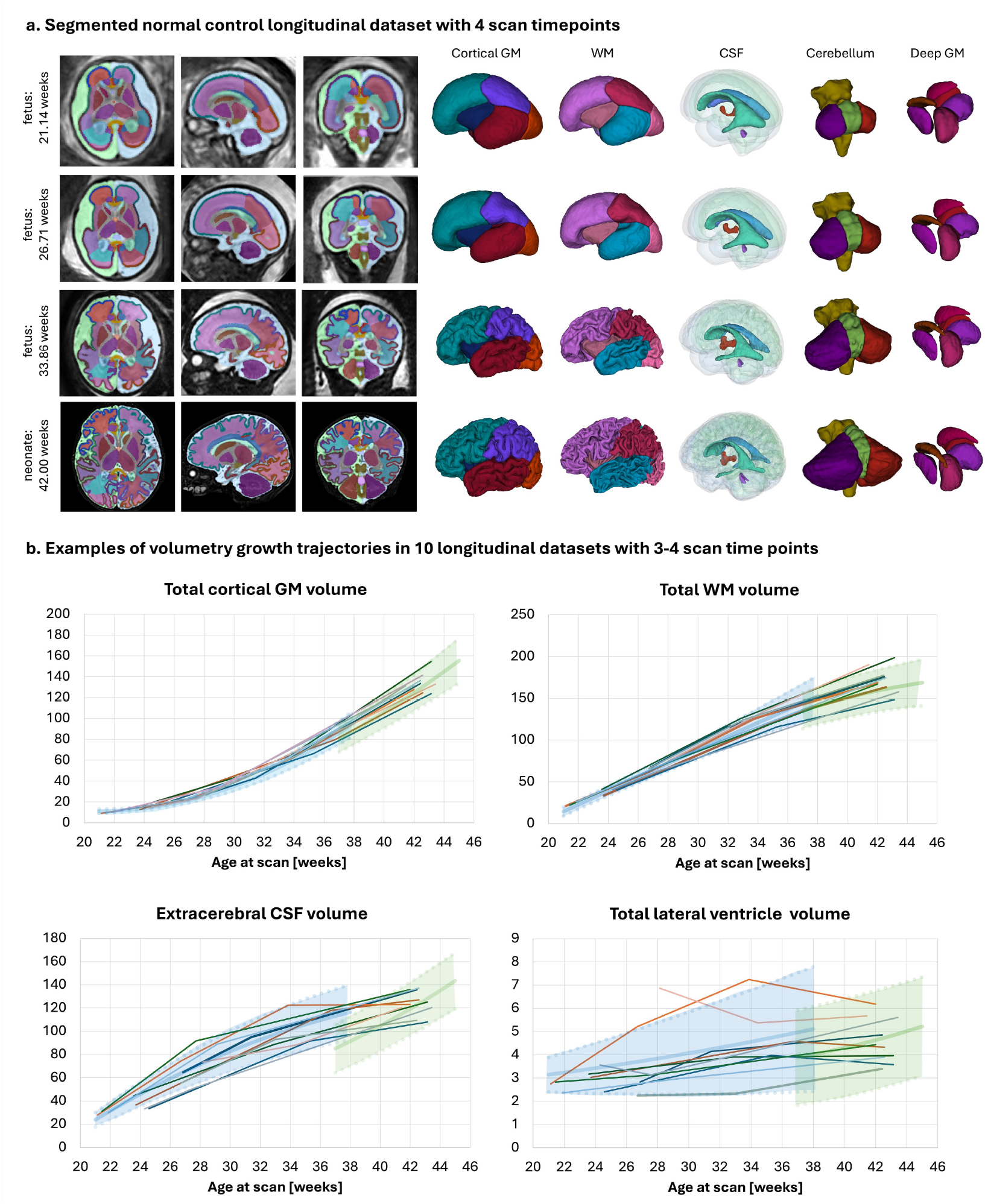
Multi-BOUNTI segmentations (a) and examples of extracted metrics (b) for 10 longitudinal datasets with more than two fetal and one neonatal scan timepoints.

### 3.4. Automated reporting

An example .html volumetry report generated from Multi-BOUNTI segmentation of a preterm neonatal dataset (GA at birth 23.85 weeks; PMA at scan 41.86 weeks) with confirmed clinically significant abnormalities is shown in Fig. 11. Although total brain, WM and cortical GM volumes fall within normal ranges, centile analysis indicates that extracerebral CSF, both lateral ventricles and the third ventricle are above the 95th centile, while posterior fossa structures and thalamus are below the 5th centile.

**Figure 11:**
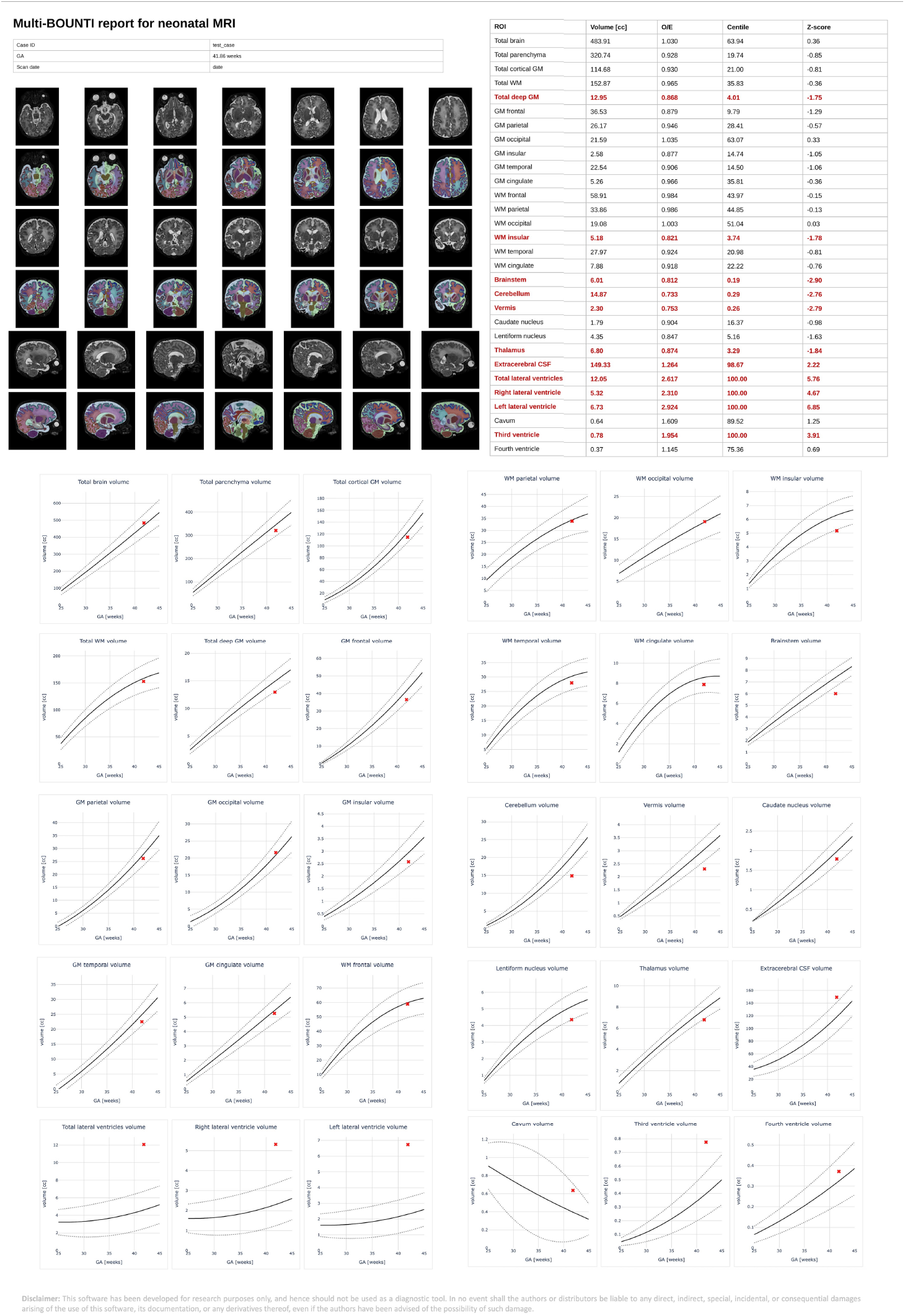
Example of an automated HTML volumetry report generated from neonatal Multi-BOUNTI segmentation.

The report format and centile-based interpretation were reviewed by expert perinatal neuroradiologists and clinicians and considered clear and meaningful for full quantitative volumetric assessment of the brain.

### 3.5. Initial evaluation of generalisation and anomaly handling

Qualitative assessment of the 405 iFIND fetal MRI reconstructions (Fig. 12) showed that Multi-BOUNTI generally achieved good segmentation quality across the gestational age range (21–38 weeks), with most cases assigned the highest quality grade (grade 4). Cases were concentrated between 22 and 34 weeks, with the largest numbers at 31–32 weeks. Minor errors (grade 3) were observed in a subset of cases, while significant errors (grade 2) and severe errors (grade 1) were uncommon. Representative examples illustrate the grading criteria, ranging from substantial segmentation inaccuracies to good anatomical delineation. Overall, these findings demonstrate promising generalisability to fetal MRI acquired at a different field strength and using a different acquisition protocol, without dedicated segmentation model retraining, although performance remains dependent on image quality and anatomical presentation.

**Figure 12:**
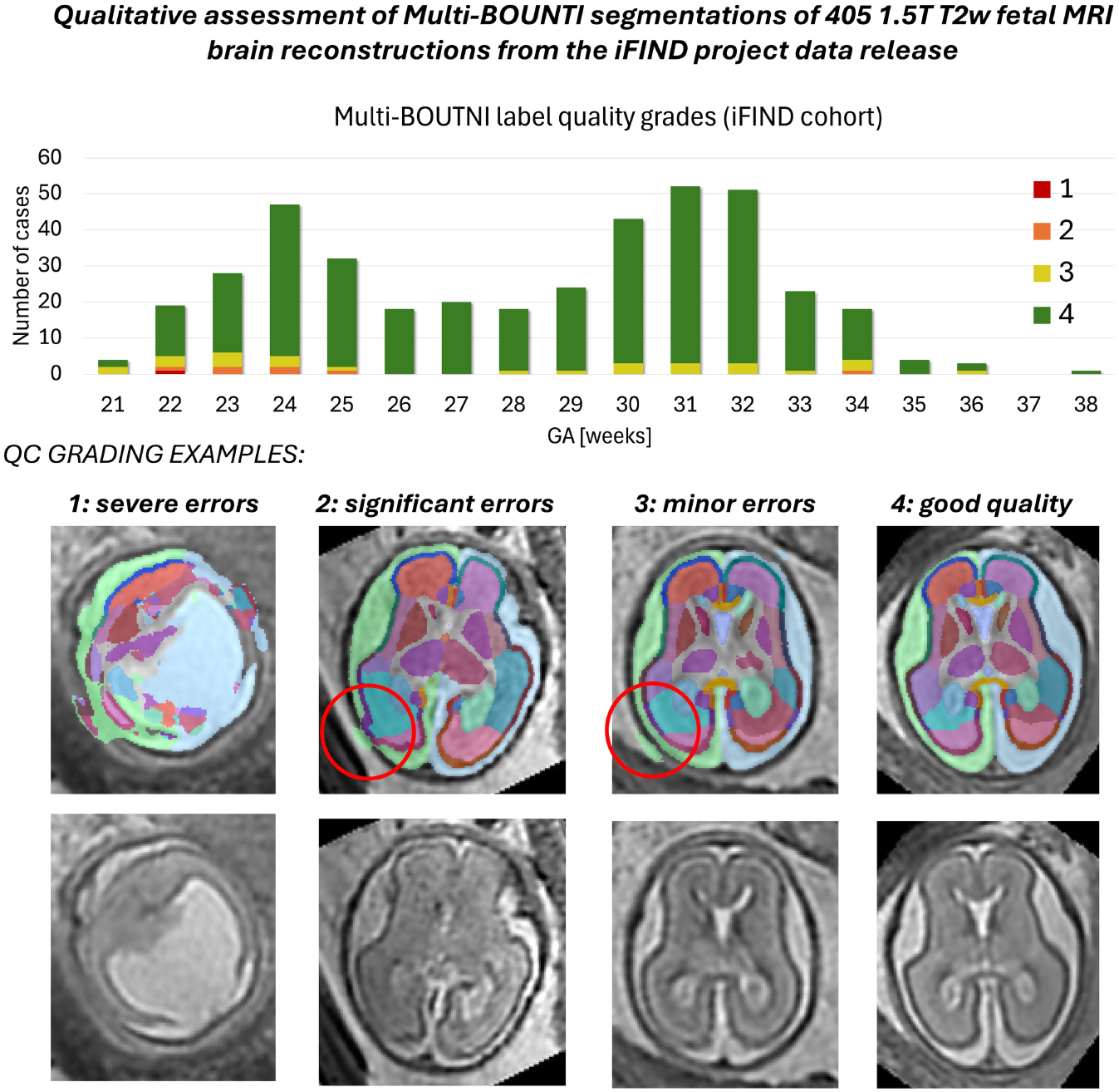
Quality assessment of Multi-BOUNTI segmentations on 405 1.5T fetal brain MRI reconstructions from iFIND project cohort.

Fig. 13 shows representative examples of MultiBOUNTI performance across various types of fetal and neonatal brain anomalies. These examples illustrate the ability of Multi-BOUNTI to delineate major brain structures in the presence of anatomical deviations, including ventricular enlargement, abnormal cortical development and focal tissue abnormalities. While the segmentations generally preserve the anatomical organisation of major brain regions, local inaccuracies are observed in areas affected by pronounced structural abnormalities or altered tissue appearance. These examples illustrate the potential of Multi-BOUNTI for analysing abnormal fetal and neonatal brain MRI, while highlighting limitations in regions with atypical anatomy.

**Figure 13:**
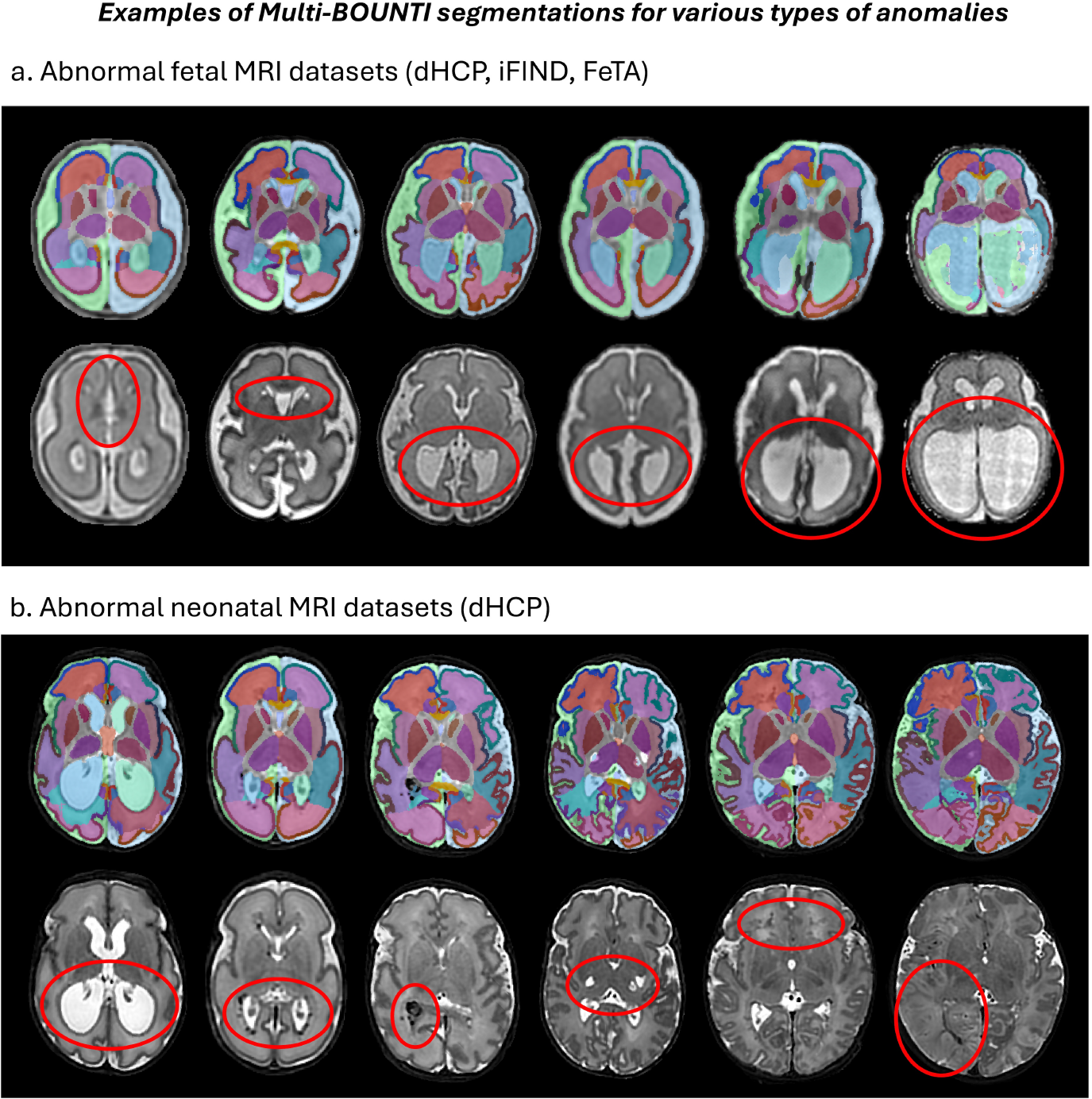
Representative examples of Multi-BOUNTI segmentations in fetal (a) and neonatal (b) MRI datasets with congenital brain malformations and acquired brain injuries. Red circles highlight regions with abnormal anatomical or tissue appearances.

## 4. Discussion

This work presents a deep learning framework that allows automated multi-lobe segmentation of T2w perinatal brain MRI across both fetal and neonatal periods. It builds on the Draw-EM pipeline (Makropoulos et al., 2018) and the large dHCP cohort of T2w datasets (Edwards et al., 2022), providing fast, robust segmentation and integrated normative volumetry modelling.

To ensure clinical and research relevance under realistic imaging constraints, we developed a dedicated parcellation protocol comprising 43 anatomical ROIs through consensus between neuroradiologists and imaging scientists. Based on this protocol, the Multi-BOUNTI pipeline enables automated segmentation of fetal and neonatal T2w MRI and generation of volumetric reports.

The segmentation framework combines automated preprocessing (brain localisation and standard space alignment) with a 3D Attention UNet trained on 160 fetal and neonatal datasets. Ground truth labels were generated using an iterative active learning strategy with manual refinement. Evaluation on 40 independent datasets (20–45 weeks age at scan) demonstrated consistently high agreement with manually refined segmentations, with relative volumetric differences below 1% and high qualitative scores across the 43 labels. The pipeline is publicly available (https://github.com/SVRTK/perinatal-brain-mri-analysis) and processes each case in under 10 minutes, supporting large-scale applications and suitability for clinical reporting timeframes.

The pipeline was further applied to 267 fetal and 494 term and 99 preterm neonatal datasets without reported clinically significant abnormalities across 21–44 weeks GA/PMA. Extracted volumetric measures showed expected developmental trajectories, with strong associations with age at scan (*p* < 0.0001), and additional associations with sex and birth weight centile (neonates). No significant association with mode of delivery was observed after correction for multiple comparisons. Due to the imbalance in ethnic representation across cohorts, interpretation of the impact of parental ethnicity was inherently limited. Lobe-specific analysis of GM, WM, cortical surface area and gyrification index demonstrated preserved inter-regional relationships across development and the fetal–neonatal transition, consistent with findings from longitudinal analysis.

These data were used to derive normative volumetric growth models for fetal and neonatal brain development, enabling computation of subject-specific centiles and zscores within an automated reporting framework. The resulting .html reports, incorporating segmentation overlays, tabulated measurements and growth charts, were reviewed by expert perinatal neuroradiologists and considered clinically relevant. This supports the feasibility of integrating 3D segmentation-derived metrics into routine radiological reporting workflows in addition to standard 2D biometry. Clinical significance of these improved phenotypes would need to be determined in further studies. The reported centiles and z-scores provide age-specific reference measurements rather than diagnostic classification. Furthermore, the normative models are specific to the Multi-BOUNTI parcellation protocol, and measurements derived using other segmentation methods may not be directly comparable.

Overall, the proposed Multi-BOUNTI pipeline provides a scalable baseline framework for unified fetal and neonatal brain segmentation and quantitative analysis, supporting both large-scale research studies and clinically interpretable volumetric assessment.

### 4.1. Limitations and future work

A key limitation is that training was performed exclusively on dHCP datasets acquired on a 3T scanner with specific acquisition parameters (e.g., TE=250 ms for fetal MRI), high image quality, and predominantly typically developing anatomy. Nevertheless, qualitative evaluation on 405 independent 1.5T fetal MRI reconstructions from the iFIND cohort demonstrated generally good segmentation quality, suggesting promising generalisability across acquisition protocols without dedicated retraining of the segmentation model. Further generalisation will require systematic validation across a wider range of acquisition protocols and image qualities. This will necessitate expansion of the training cohort to include more heterogeneous datasets with possible application of harmonisation (Grigorescu et al., 2021) and synthetic learning (Billot et al., 2023; Valabregue et al., 2024) strategies.

Although a subset of abnormal cases from the dHCP cohort was included in training, this was not systematic and does not cover the full spectrum of fetal and neonatal brain anomalies. Nevertheless, representative examples of congenital malformations and acquired brain injuries showed that Multi-BOUNTI can delineate major anatomical structures across a range of abnormal presentations, suggesting potential applicability to pathological datasets. However, these initial observations require further systematic validation and dedicated retraining using clinically representative pathological datasets to improve robustness across different types and severities of abnormalities.

Importantly, segmentation performance is inherently dependent on input image quality. Severe motion artefacts, low signal-to-noise ratio, reconstruction errors and poorly defined anatomical boundaries may intrinsically limit segmentation accuracy, independently of model performance. These image-quality-related limitations should be distinguished from model-specific segmentation errors. This highlights the importance of using highquality input reconstructions and applying appropriate image quality assessment before segmentation and downstream volumetric analysis. Automated quality control for both input images (Sanchez et al., 2024) and output segmentations will also be essential to minimise propagation of errors and uncertainty into downstream analyses. The current framework requires reconstructed 3D T2w images with sufficient anatomical resolution and contrast and is not designed for direct segmentation of individual 2D acquisition stacks. Application to lower-resolution clinical reconstructions will require further validation.

The parcellation protocol will be further extended to include additional structures relevant to brain development, such as the hippocampus, amygdala, midbrain, pons, nucleus accumbens, hypothalamus, choroid plexus and internal capsule, as well as finer sub-parcellation of cortical GM/WM (e.g., motor cortex). It will also require adaptation to accommodate pathological cases with altered anatomy or the presence of additional structures (e.g., cystic lesions, tumours or haemorrhages).

Future work will also focus on improving normative modelling by incorporating Gaussian process approaches (Dimitrova et al., 2021) and subject-specific multi-factor modelling (e.g., sex, birth weight, fetal size), alongside more detailed consideration of clinical metadata in cohort selection. We will also increase the cohort of longitudinal scans across the perinatal cohort to improve our understanding of brain development.

The current results also suggest the feasibility of further extension of the subject age range to infancy and other MRI contrasts.

Finally, given the improved cortical delineation achieved by Multi-BOUNTI, extending the framework to cortical surface reconstruction and combined fetal–neonatal surface analysis represents a natural next step, building on previous work that used tissue-only BOUNTI (Uus et al., 2023) for surface extraction (Sanchez et al., 2026; Grigorescu et al., 2026).

## 5. Conclusions

We present Multi-BOUNTI, a unified deep learning framework for automated multi-lobe segmentation of fetal and neonatal T2w brain MRI. The approach enables consistent parcellation across the perinatal period and provides fast, robust segmentation.

Applied to a large dHCP cohort, the pipeline enables derivation of normative volumetric growth models and supports subject-specific assessment via centiles and z-scores within an automated reporting framework.

Overall, Multi-BOUNTI provides a scalable baseline solution for combined fetal and neonatal brain segmentation and volumetry, with potential for both research and clinical applications.

## Data Availability

The MRI datasets used in this work are publicly available at: https://www.developingconnectome.org/data-release

## Acknowledgments

We thank everyone who was involved in the acquisition and analysis of the datasets at the Department of Early Life Imaging at King’s College London. We thank all participants and families.

This work was supported by Rosetrees Trust awarded to MR [2817415], NIHR Advanced Fellowship awarded to LS [NIHR30166], AMR GN4028 awarded to MR [3788508], the European Research Council under the European Union’s Seventh Framework Programme [FP7/20072013]/ERC grant agreement no. 319456 dHCP project, the BIBS project has received funding from the Innovative Medicines Initiative 2 Joint Undertak- ing under grant agreement No 777394, the Wellcome/EPSRC Centre for Medical Engineering at King’s College London [WT 203148/Z/16/Z], MRC Centre for Neurodevelopmental Disorders King’s College London [MR/N026063/1], the NIHR Clinical Research Facility (CRF) at Guy’s and St Thomas’ and by the National Institute for Health Research Biomedical Research Centre based at Guy’s and St Thomas’ NHS Foundation Trust and King’s College London. TA is funded by an MRC Senior Clinical Fellowship [MR/Y009665/1].

ChatGPT (OpenAI) was used to assist with language editing and refinement of the manuscript. All scientific content was reviewed and verified by the authors, who take full responsibility for the final manuscript.

The views expressed are those of the authors and not necessarily those of the NHS, the NIHR or the Department of Health.

## Author contributions

AU developed the segmentation pipeline, performed data processing and analysis, and prepared the manuscript. AFG contributed to the design of the parcellation protocol, literature review, data processing, and qualitative evaluation of the datasets. VK contributed to the parcellation protocol design, data processing, and analysis. DC, TM, SA, AL, HS, RB, CB and AEC contributed to the development of the parcellation protocol. AM, AS, ER, MD, LC and JM contributed to processing of the dHCP datasets. AP, JH and KC contributed to acquisition of the dHCP datasets. AH contributed to the initial parcellation protocol design. DR, SC, GM and TA supervised methodological components related to dHCP data processing and analysis. ADE and JH provided access to the dHCP datasets and supervised aspects of the project. MR contributed to the parcellation protocol design and radiological assessment of the datasets and supervised the project. LS supervised the project. All authors reviewed and approved the manuscript.

## Footnotes

1 MIRTK toolbox: https://github.com/biomedia/mirtk

## References

Adamson, C.L., Alexander, B., Kelly, C.E., Ball, G., Beare, R., Cheong, J.L., Spittle, A.J., Doyle, L.W., Anderson, P.J., Seal, M.L., Thompson, D.K., 2024. Updates to the melbourne children’s regional infant brain software package (m-crib-s). Neuroinformatics 22, 207–223. doi:10.1007/s12021-024-09656-8.

Bagheri, M., Velasco-Annis, C., Wang, J., Faghihpirayesh, R., Khan, S., Calixto, C., Jaimes, C., Vasung, L., Ouaalam, A., Afacan, O., Warfield, S.K., Rollins, C.K., Gholipour, A., 2026. An mri atlas of the human fetal brain: Reference and segmentation tools for fetal brain mri analysis. doi:10.48550/arXiv.2508.15034, arXiv:2508.15034.

Ball, G., Aljabar, P., Nongena, P., Kennea, N., Gonzalez-Cinca, N., Falconer, S., Chew, A.T., Harper, N., Wurie, J., Rutherford, M.A., Counsell, S.J., Edwards, A.D., 2017. Multimodal image analysis of clinical influences on preterm brain development. Annals of Neurology 82, 233–246. doi:10.1002/ana.24995.

Bayer, S., Altman, J., 2003. The Human Brain During the Third Trimester. United Kingdom: CRC Press. doi:10.1201/9780203494943.

Bayer, S., Altman, J., 2005. The Human Brain During the Second Trimester. United Kingdom: CRC Press. doi:10.1201/9780203507483.

Billot, B., Greve, D.N., Puonti, O., Thielscher, A., Leemput, K.V., Fischl, B., Dalca, A.V., Iglesias, J.E., 2023. Synthseg: Segmentation of brain mri scans of any contrast and resolution without retraining. Medical Image Analysis 86. doi:10.1016/j.media.2023.102789.

Cardoso, M.J., Li, W., Brown, R., Ma, N., Kerfoot, E., Wang, Y., Murrey, B., Myronenko, A., Zhao, C., Yang, D., Nath, V., He, Y., Xu, Z., Hatamizadeh, A., Myronenko, A., Zhu, W., Liu, Y., Zheng, M., Tang, Y., Yang, I., Zephyr, M., Hashemian, B., Alle, S., Darestani, M.Z., Budd, C., Modat, M., Vercauteren, T., Wang, G., Li, Y., Hu, Y., Fu, Y., Gorman, B., Johnson, H., Genereaux, B., Erdal, B.S., Gupta, V., Diaz-Pinto, A., Dourson, A., Maier-Hein, L., Jaeger, P.F., Baumgartner, M., Kalpathy-Cramer, J., Flores, M., Kirby, J., Cooper, L.A.D., Roth, H.R., Xu, D., Bericat, D., Floca, R., Zhou, S.K., Shuaib, H., Farahani, K., Maier-Hein, K.H., Aylward, S., Dogra, P., Ourselin, S., Feng, A., 2022. Monai: An open-source framework for deep learning in healthcare. ArXiv doi:10.48550/arXiv.2211.02701.

Cicek, O., Abdulkadir, A., Lienkamp, S.S., Brox, T., Ronneberger, O., 2016. 3d u-net: Learning dense volumetric segmentation from sparse annotation. ArXiv abs/1606.06650. doi:10.48550/arXiv.1606.06650.

Cordero-grande, L., Price, A.N., Hughes, E.J., Wright, R., Rutherford, M.A., Hajnal, J.V., 2019. Automating fetal brain reconstruction using distance regression learning, in: ISMRM, pp. 3–6. URL: https://archive.ismrm.org/2019/4779.html.

Cromb, D., Uus, A., Poppel, M.P.M.V., Steinweg, J.K., Bonthrone, A.F., Maggioni, A., Cawley, P., Egloff, A., Kyriakopolous, V., Matthew, J., Price, A., Pushparajah, K., Simpson, J., Razavi, R., DePrez, M., Edwards, D., Hajnal, J., Rutherford, M., Lloyd, D.F.A., Counsell, S.J., 2024. Total and regional brain volumes in fetuses with congenital heart disease. Journal of Magnetic Resonance Imaging 60, 497–509. doi:10.1002/jmri.29078.

Cuadra, M.B., Sanchez, T., Juan, G.M., Auzias, G., 2025. Protocol for the quality rating of 3D super-resolution reconstruction in fetal brain MRI. Technical Report. Zenodo. doi:10.5281/zenodo.15696638.

Deprest, T., Fidon, L., Keyzer, F.D., Ebner, M., Deprest, J., Demaerel, P., Catte, L.D., Vercauteren, T., Ourselin, S., Dymarkowski, S., Aertsen, M., 2023. Application of automatic segmentation on super-resolution reconstruction mr images of the abnormal fetal brain. American Journal of Neuroradiology 44, 486–491. doi:10.3174/ajnr.A7808.

Dimitrova, R., Arulkumaran, S., Carney, O., Chew, A., Falconer, S., Ciarrusta, J., Wolfers, T., Batalle, D., Cordero-Grande, L., Price, A.N., Teixeira, R.P.A.G., Hughes, E., Egloff, A., Hutter, J., Makropoulos, A., Robinson, E.C., Schuh, A., Vecchiato, K., Steinweg, J.K., Macleod, R., Marquand, A.F., McAlonan, G., Rutherford, M.A., Counsell, S.J., Smith, S.M., Rueckert, D., Hajnal, J.V., O’Muircheartaigh, J., Edwards, A.D., 2021. Phenotyping the preterm brain: Characterizing individual deviations from normative volumetric development in two large infant cohorts. Cerebral Cortex 31, 3665–3677. doi:10.1093/cercor/bhab039.

Edwards, A.D., Rueckert, D., Smith, S.M., Seada, S.A., Alansary, A., Almalbis, J., Allsop, J., Andersson, J., Arichi, T., Arulkumaran, S., Bastiani, M., Batalle, D., Baxter, L., Bozek, J., Braithwaite, E., Brandon, J., Carney, O., Chew, A., Christiaens, D., Chung, R., Colford, K., Cordero-Grande, L., Counsell, S.J., Cullen, H., Cupitt, J., Curtis, C., Davidson, A., Deprez, M., Dillon, L., Dimitrakopoulou, K., Dimitrova, R., Duff, E., Falconer, S., Farahibozorg, S.R., Fitzgibbon, S.P., Gao, J., Gaspar, A., Harper, N., Harrison, S.J., Hughes, E.J., Hutter, J., Jenkinson, M., Jbabdi, S., Jones, E., Karolis, V., Kyriakopoulou, V., Lenz, G., Makropoulos, A., Malik, S., Mason, L., Mortari, F., Nosarti, C., Nunes, R.G., O’Keeffe, C., O’Muircheartaigh, J., Patel, H., PasseratPalmbach, J., Pietsch, M., Price, A.N., Robinson, E.C., Rutherford, M.A., Schuh, A., Sotiropoulos, S., Steinweg, J., Teixeira, R.P.A.G., Tenev, T., Tournier, J.D., Tusor, N., Uus, A., Vecchiato, K., Williams, L.Z.J., Wright, R., Wurie, J., Hajnal, J.V., 2022. The developing human connectome project neonatal data release. Frontiers in Neuroscience 16. doi:10.3389/fnins.2022.886772.

Fidon, L., Aertsen, M., Kofler, F., Bink, A., David, A.L., Deprest, T., Emam, D., Guffens, F., Jakab, A., Kasprian, G., Kienast, P., Melbourne, A., Menze, B., Mufti, N., Pogledic, I., Prayer, D., Stuempflen, M., Elslander, E.V., Ourselin, S., Deprest, J., Vercauteren, T., 2024. A dempster-shafer approach to trustworthy ai with application to fetal brain mri segmentation. arXiv arXiv:2204.02779. doi:10.1109/TPAMI.2023.3346330.

Gholipour, A., Rollins, C.K., Velasco-Annis, C., Ouaalam, A., Akhondi-Asl, A., Afacan, O., Ortinau, C.M., Clancy, S., Limperopoulos, C., Yang, E., Estroff, J.A., Warfield, S.K., 2017. A normative spatiotemporal mri atlas of the fetal brain for automatic segmentation and analysis of early brain growth. Nature: Scientific Reports 7, 1–13. doi:10.1038/s41598-017-00525-w.

Gousias, I.S., Edwards, A.D., Rutherford, M.A., Counsell, S.J., Hajnal, J.V., Rueckert, D., Hammers, A., 2012. Magnetic resonance imaging of the newborn brain: Manual segmentation of labelled atlases in term-born and preterm infants. NeuroImage 62, 1499–1509. doi:10.1016/j.neuroimage.2012.05.083.

Grigorescu, I., Vanes, L., Uus, A., Batalle, D., Cordero-Grande, L., Nosarti, C., Edwards, A.D., Hajnal, J.V., Modat, M., Deprez, M., 2021. Harmonized segmentation of neonatal brain mri. Frontiers in Neuroscience 15. doi:10.3389/fnins.2021.662005.

Grigorescu, I., Xiao, J., Guo, Y., Kyriakopoulou, V., Uus, A., Karolis, V., Liang, K., Suliman, M.A., Ma, Q., Rueckert, D., Kainz, B., Edwards, A.D., Hajnal, J., Rutherford, M., Deprez, M., Robinson, E.C., 2026. Sud-coTAN: Sulcal depth-guided anatomically consistent fetal cortical surface reconstruction, in: Medical Imaging with Deep Learning. URL: https://openreview.net/forum?id=1ZKHyRKUW9.

Hendrickson, T.J., Reiners, P., Moore, L.A., Lundquist, J.T., Fayzullobekova, B., Perrone, A.J., Lee, E.G., Moser, J., Day, T.K.M., Alexopoulos, D., Styner, M., Kardan, O., Chamberlain, T.A., Mummaneni, A., Caldas, H.A., Bower, B., Stoyell, S., Martin, T., Sung, S., Fair, E.A., Carter, K., Uriarte-Lopez, J., Rueter, A.R., Yacoub, E., Rosenberg, M.D., Smyser, C.D., Elison, J.T., Graham, A., Fair, D.A., Feczko, E., 2026. Bibsnet: A deep learning baby image brain segmentation network for mri scans. Developmental Cognitive Neuroscience 79, 101706. doi:10.1016/j.dcn.2026.101706.

Henschel, L., Kügler, D., Zöllei, L., Reuter, M., 2024. Vinna for neonates: Orientation independence through latent augmentations. Imaging Neuroscience 2, 1–26. doi:10.1162/imag_a_00180.

Huang, S., Zhang, K., Zhu, F., Ding, Z., Chen, G., Shen, D., 2026. Semi-supervised fetal brain parcellation via hierarchical learning framework. Medical Image Analysis 107, 103835. doi:10.1016/j.media.2025.103835.

Isensee, F., Jaeger, P.F., Kohl, S.A., Petersen, J., Maier-Hein, K.H., 2021. nnu-net: a self-configuring method for deep learning-based biomedical image segmentation. Nature Methods 18, 203–211. doi:10.1038/s41592-020-01008-z.

Khan, Y.T., Tsompanidis, A., Radecki, M.A., Allison, C., Lai, M.C., Bethlehem, R.A., Baron-Cohen, S., 2026. Mapping brain growth and sex differences across prenatal to postnatal development. Scientific Reports 16. doi:10.1038/s41598-025-33981-w.

Kuklisova-Murgasova, M., Quaghebeur, G., Rutherford, M.A., Hajnal, J.V., Schnabel, J.A., 2012. Reconstruction of fetal brain mri with intensity matching and complete outlier removal. Medical Image Analysis 16, 1550–1564. doi:10.1016/j.media.2012.07.004.

Lefévre, J., Germanaud, D., Dubois, J., Rousseau, F., Santos, I.D.M., Angleys, H., Mangin, J.F., Hüppi, P.S., Girard, N., Guio, F.D., 2016. Are developmental trajectories of cortical folding comparable between cross-sectional datasets of fetuses and preterm new-borns? Cerebral Cortex 26, 3023–3035. doi:10.1093/cercor/bhv123.

Ma, Q., Liang, K., Li, L., Masui, S., Guo, Y., Nosarti, C., Robinson, E.C., Kainz, B., Rueckert, D., 2025. The developing human connectome project: A fast deep learning-based pipeline for neonatal cortical surface reconstruction. Medical Image Analysis 100, 103394. doi:10.1016/j.media.2024.103394.

Makropoulos, A., Robinson, E.C., Schuh, A., Wright, R., Fitzgibbon, S., Bozek, J., Counsell, S.J., Steinweg, J., Vecchiato, K., Passerat-Palmbach, J., Lenz, G., Mortari, F., Tenev, T., Duff, E.P., Bastiani, M., Cordero-Grande, L., Hughes, E., Tusor, N., Tournier, J.D., Hutter, J., Price, A.N., Teixeira, R.P.A.G., Murgasova, M., Victor, S., Kelly, C., Rutherford, M.A., Smith, S.M., Edwards, A.D., Hajnal, J.V., Jenkinson, M., Rueckert, D., 2018. The developing human connectome project: A minimal processing pipeline for neonatal cortical surface reconstruction. NeuroImage 173, 88–112. doi:10.1016/j.neuroimage.2018.01.054.

Mihailov, A., Pron, A., Lefévre, J., Deruelle, C., Desnous, B., Bretelle, F., Manchon, A., Milh, M., Rousseau, F., Girard, N., Auzias, G., 2025. Burst of gyrification in the human brain after birth. Communications Biology 8, 805. doi:10.1038/s42003-025-08155-z.

Modi, N., Bétrémieux, P., Midgley, J., Hartnoll, G., 2000. Postnatal weight loss and contraction of the extracellular compartment is triggered by atrial natriuretic peptide. Early Human Development 59, 201–208. doi:10.1016/S0378-3782(00)00097-9.

National Guideline Alliance (UK), 2017. Weight loss in the early days of life, in: Faltering Growth: Recognition and Management. National Institute for Health and Care Excellence (NICE), London. NICE Guideline, No. 75. chapter 4. URL: https://www.ncbi.nlm.nih.gov/books/NBK536449/.

Oktay, O., Schlemper, J., Folgoc, L., Lee, M., Heinrich, M., Misawa, K., Mori, K., McDonagh, S., Hammerla, N., Kainz, B., Glocker, B., Rueckert, D., 2018. Attention u-net: Learning where to look for the pancreas. ArXiv arXiv:1804.03999. doi:10.48550/arXiv.1804.03999.

Orasanu, E., Melbourne, A., Cardoso, M.J., Lomabert, H., Kendall, G.S., Robertson, N.J., Marlow, N., Ourselin, S., 2016. Cortical folding of the preterm brain: a longitudinal analysis of extremely preterm born neonates using spectral matching. Brain and Behavior 6. doi:10.1002/brb3.488.

Payette, K., Steger, C., Licandro, R., d. Dumast, P., Li, H.B., Barkovich, M., Li, L., Dannecker, M., Chen, C., Ouyang, C., McConnell, N., Miron, A., Li, Y., Uus, A., Grigorescu, I., Gilliland, P.R., Siddiquee, M.M.R., Xu, D., Myronenko, A., Wang, H., Huang, Z., Ye, J., Alenyá, M., Comte, V., Camara, O., Masson, J.B., Nilsson, A., Godard, C., Mazher, M., Qayyum, A., Gao, Y., Zhou, H., Gao, S., Fu, J., Dong, G., Wang, G., Rieu, Z., Yang, H., Lee, M., Płotka, S., Grzeszczyk, M.K., Sitek, A., Daza, L.V., Usma, S., Arbelaez, P., Lu, W., Zhang, W., Liang, J., Valabregue, R., Joshi, A.A., Nayak, K.N., Leahy, R.M., Wilhelmi, L., Dändliker, A., Ji, H., Gennari, A.G., Jakovčić, A., Klaić, M., Adžić, A., Marković, P., Grabarić, G., Kasprian, G., Dovjak, G., Rados, M., Vasung, L., Cuadra, M.B., Jakab, A., 2025a. Multi-center fetal brain tissue annotation (feta) challenge 2022 results. IEEE Transactions on Medical Imaging 44, 1257–1272. doi:10.1109/TMI.2024.3485554.

Payette, K., Uus, A.U., Kollstad, E., Verdera, J.A., Gallo, D., Hall, M., Hajnal, J.V., Rutherford, M.A., Story, L., Hutter, J., 2025b. T2* relaxometry of fetal brain structures using low-field (0.55t) mri. Magnetic Resonance in Medicine 93, 1942–1953. doi:10.1002/mrm.30409.

Price, A.N., Cordero-grande, L., Hughes, E., Hiscocks, S., Green, E., Mccabe, L., Ferrazzi, G., Deprez, M., Roberts, T., Christiaens, D., Duff, E., Karolis, V., Malik, J., Rutherford, M.A., Edwards, D.A., Hajnal, J.V., 2019. The developing human connectome project (dhcp): fetal acquisition protocol, in: ISMRM. URL: https://archive.ismrm.org/2019/0244.html.

Richter, L., Fetit, A.E., 2022. Accurate segmentation of neonatal brain mri with deep learning. Frontiers in Neuroinformatics Volume 16.

Royston, P., Wright, E., 1998. How to construct ‘normal ranges’ for fetal variables. Ultrasound in Obstetrics & Gynecology 11, 30–38. doi:10.1046/j.1469-0705.1998.11010030.x.

Sanchez, T., Esteban, O., Gomez, Y., Pron, A., Koob, M., Dunet, V., Girard, N., Jakab, A., Eixarch, E., Auzias, G., Cuadra, M.B., 2024. Fetmrqc: A robust quality control system for multi-centric fetal brain mri. Medical Image Analysis, 103282 doi: 10.1016/j.media.2024.103282.

Sanchez, T., Martí-Juan, G., Meunier, D., Ballester, M.A.G., Camara, O., Eixarch, E., Piella, G., Cuadra, M.B., Auzias, G., 2026. Fetpype: An open-source pipeline for reproducible fetal brain mri analysis. ArXiv doi:10.48550/arXiv.2512.17472.

Story, L., Davidson, A., Patkee, P., Fleiss, B., Kyriakopoulou, V., Colford, K., Sankaran, S., Seed, P., Jones, A., Hutter, J., Shennan, A., Rutherford, M., 2021. Brain volumetry in fetuses that deliver very preterm: An mri pilot study. NeuroImage: Clinical 30, 102650. doi:10.1016/j.nicl.2021.102650.

Tustison, N.J., Gee, J.C., 2009. N4itk: Nick’s n3 itk implementation for mri bias field correction. InsightJournal, 1–8doi:10.54294/jculxw.

Urru, A., Benkarim, O., Martí-Juan, G., Hahner, N., Piella, G., Eixarch, E., Ballester, M.A.G., 2025. Longitudinal assessment of abnormal cortical folding in fetuses and neonates with isolated non-severe ventriculomegaly. Brain and Behavior 15, e70255. doi:10.1002/brb3.70255.

Uus, A., Silva, S.N., Verdera, J.A., Payette, K., Hall, M., Colford, K., Luis, A., Sousa, H., Ning, Z., Roberts, T., McElroy, S., Deprez, M., Hajnal, J., Rutherford, M., Story, L., Hutter, J., 2025. Scanner-based real-time three-dimensional brain+body slice-to-volume reconstruction for t2-weighted 0.55-t low-field fetal magnetic resonance imaging. Pediatric Radiology doi:10.1007/s00247-025-06165-x.

Uus, A.U., Collado, A.E., Roberts, T.A., Hajnal, J.V., Rutherford, M.A., Deprez, M., 2022. Retrospective motion correction in foetal mri for clinical applications: existing methods, applications and integration into clinical practice. The British Journal of Radiology doi:10.1259/bjr.20220071.

Uus, A.U., Kyriakopoulou, V., Makropoulos, A., Fukami-Gartner, A., Cromb, D., Davidson, A., Cordero-Grande, L., Price, A.N., Grigorescu, I., Williams, L.Z.J., Robinson, E.C., Lloyd, D., Pushparajah, K., Story, L., Hutter, J., Counsell, S.J., Edwards, A.D., Rutherford, M.A., Hajnal, J.V., Deprez, M., 2023. Bounti: Brain volumetry and automated parcellation for 3d fetal mri. eLife 12:RP88818. doi:10.7554/elife.88818.1.

Valabregue, R., Girka, F., Pron, A., Rousseau, F., Auzias, G., 2024. Comprehensive analysis of synthetic learning applied to neonatal brain mri segmentation. Human Brain Mapping 45. doi:10.1002/hbm.26674.

Xu, X., Sun, C., Sun, J., Shi, W., Shen, Y., Zhao, R., Luo, W., Li, M., Wang, G., Wu, D., 2022. Spatiotemporal atlas of the fetal brain depicts cortical developmental gradient. Journal of Neuroscience 42, 9435–9449. doi:10.1523/JNEUROSCI.1285-22.2022.

Yushkevich, P.A., Piven, J., Hazlett, H.C., Smith, R.G., Ho, S., Gee, J.C., Gerig, G., 2006. User-guided 3d active contour segmentation of anatomical structures: Significantly improved efficiency and reliability. NeuroImage 31, 1116–1128. doi:10.1016/j.neuroimage.2006.01.015.

Zalevskyi, V., Sanchez, T., Roulet, M., Bulut, B., Lajous, H., Verdera, J.A., Silva, S.N., Langs, G., Kasprian, G., Licandro, R., Hutter, J., Kebiri, H., Cuadra, M.B., 2026. Evaluating synthetic data generation for domain generalization in fetal brain mri segmentation. Machine Learning for Biomedical Imaging 2026, 482–506. doi:10.59275/j.melba.2026-6e4e.

Zeng, Q., Liu, W., Li, B., Didier, R., Grant, P.E., Karimi, D., 2026. Atlas-assisted segment anything model for fetal brain mri (fetal-sam). doi:10.48550/arXiv.2601.15759, arXiv:2601.15759.

Zilles, K., Armstrong, E., Schleicher, A., Kretschmann, H.J., 1988. The human pattern of gyrification in the cerebral cortex. Anatomy and Embryology 179, 173–179. doi:10.1007/BF00304699.

